# Post-operative morbidity and mortality in Indigenous Peoples: A scoping review and meta-analysis

**DOI:** 10.1101/2023.03.27.23287510

**Authors:** Rachel J Livergant, Kelsey Stefanyk, Catherine Binda, Georgia Fraulin, Sasha Maleki, Sarah Sibbeston, Shahrzad Joharifard, Tracey Hillier, Emilie Joos

## Abstract

Indigenous Peoples across North America and Oceania experience worse health outcomes compared to non-Indigenous people, including increased post-operative morbidity and mortality. No data is available on global differences in surgical morbidity and mortality between geographic locations and across surgical specialties. The aim of this study is to evaluate disparities in post-operative morbidity and mortality between Indigenous and non-Indigenous populations. This scoping review and meta-analysis was conducted in accordance with PRISMA-ScR and MOOSE guidelines. Eight electronic databases were searched with no language restriction. Studies reporting on Indigenous populations outside of Canada, the USA, New Zealand, or Australia, or on interventional procedures were excluded. Primary outcomes were post-operative morbidity and mortality. Secondary outcomes included reoperations, readmission rates, and length of hospital stay. The Newcastle Ottawa Scale was used for quality assessment. Eighty-four unique observational studies were included in this review. Of these, 67 studies were included in the meta-analysis (Oceania n=31, North America n=36). Extensive heterogeneity existed among studies and 50% were of poor quality. Indigenous patients worldwide had 1.26 times higher odds of post-operative morbidity (OR=1.26, 95% CI: 1.10-1.44, p<0.01) and 1.34 times higher odds of post-operative infection (OR=1.34, 95% CI: 1.12-1.59, p<0.01) than non-Indigenous patients. Indigenous patients also had 1.33 times higher odds of reoperation (OR=1.33, 95% CI: 1.02-1.74, p=0.04) and longer hospital stays (SMD=0.09; 95% CI: 0.02-1.08; p=0.05). In conclusion, we found that Indigenous patients experience significantly poorer surgical outcomes than their non-Indigenous counterparts. Additionally, there remains a paucity of high-quality research focusing on assessing and improving surgical equity for Indigenous patients worldwide, despite multiple international and national calls to action for reconciliation and decolonization to improve access to quality surgical care for Indigenous populations.

## INTRODUCTION

Safe and appropriate surgical care is an integral component of an effective and resilient healthcare system [1]. Surgical conditions account for over 33% of the global disease burden [2]. Unfortunately, access to surgical care is not equitable, with populations in low-income countries, rural environments, and certain underserved populations receiving a lower quality of surgical care or no surgical care at all [1].

Indigenous Peoples are a grossly underserved population worldwide [3-5]. Although definitions of the term “Indigenous” are varied and nuanced, including populations across all continents, throughout this review we use the term Indigenous to refer to the original peoples, communities, and nations of the regions now called Canada, Australia, New Zealand, and the United States of America (USA). These populations hold and practice cultural, economic, political, and social traditions that are distinct from the broader settler population in which they live and seek recognition and sovereignty to practice these ways and identities [6]. Indigenous Peoples have not shared in the gains that development has provided these countries thus far, resulting in profound health and social inequities between Indigenous and non-Indigenous populations that continue to persist due to the ongoing effects of colonization and systemic racism [7-11].

An estimated 7 million of the 370 million Indigenous people worldwide live in Canada, Australia, New Zealand, and the USA [7]. The shared history of the British settler colonial project in these nations can be used as a framework to highlight commonalities in the contemporary situation of Indigenous Peoples in these areas [12]. For instance, Canada, Australia, New Zealand, and the USA were the only United Nations member countries to vote against The United Nations Declaration on the Rights of Indigenous Peoples (UNDRIP) upon its introduction in 2007 [13].

However, current literature on post-operative outcomes in Indigenous patients in Canada, Australia, New Zealand, and the USA, among other countries, remains limited and of poor quality [14]. Higher rates of death and adverse events post-operatively have been demonstrated for Indigenous Peoples, including increased surgical infections, need for re-operation, and longer length of hospital stay (LOS) compared to non-Indigenous patients [14-17].

To date, there is no known study that combines data on both post-operative morbidity and mortality from the continents where minority Indigenous populations experience similar disparities in healthcare provision. Additionally, no study compares and assesses morbidity and mortality for Indigenous patients across multiple surgical disciplines and procedures. This scoping review aims to assess if surgical morbidity and mortality disparities exist between Indigenous and non-Indigenous peoples in Canada, Australia, New Zealand, and the USA to better understand the extent of existing surgical and health disparities worldwide.

## METHODS

This scoping review and meta-analysis was registered in Open Science Framework (osf.io/qs3vz) and reported in accordance with Preferred Reporting Items for Systematic Review and Meta-Analysis-Extension for Scoping Reviews (PRISMA-ScR) and Meta-Analysis of Observational Studies in Epidemiology guidelines (S1 Appendix) [18,19].

### Data Sources and Searches

A search strategy was developed in consultation with a professional research librarian. Comprehensive electronic database searches were undertaken in MEDLINE, Embase, Global Health, Cochrane Library, PsycInfo, SOCIndex, Web of Science, and ProQuest Dissertations & Theses Global from inception to December 25, 2022, using key MeSH terms (S2 Appendix). All languages were included. Reference lists of reviews and retrieved articles and consultations with experts were conducted to identify additional relevant studies.

### Study Selection and Criteria

Two reviewers independently screened titles, abstracts, and full texts using Covidence. Discrepancies were resolved via consensus. We included clinical studies on surgical outcomes in Indigenous populations. Studies were excluded if they were book chapters, conference abstracts, or non-peer reviewed articles. Studies were excluded if they focused on Indigenous populations outside of Canada, Australia, New Zealand, or the USA, if they lacked a non-Indigenous comparator group, or if they included only pediatric patients, as defined in study methods. Studies describing minor interventions and procedures, such as colonoscopy or angiography, were excluded.

While we recognize the importance of accounting for the numerous and varied Indigenous populations across the world, we restricted this study’s geography to limit the already heterogeneous nature of our data. Furthermore, we hope to avoid over-homogenizing the distinct lived experiences of Indigenous Peoples in regions such as Africa, Northern Europe, and Central/South America. Canada, Australia, New Zealand and the USA share similar British colonial settler histories and consequent displacement and oppression of native people [7, 20-22].

### Data Extraction and Quality Assessment

One reviewer completed data extraction and quality assessment (QA), while another verified the extracted data and QA findings. Discrepancies were resolved through consensus. The following data were extracted from included studies using Microsoft Excel (Microsoft Corporation, Version 16.60): authors’ name, journal, year of publication, age category, population size, sex, type of study, database, surgical specialty, operations, outcomes of interest, and study conclusions. “Outcomes of interest” included mortality, any morbidity, length of hospital stay, and readmission and reoperation rates. Studies were included in data extraction if they reported surgical procedures and at least one outcome of interest resulting from the procedure. Studies reporting on two separate Indigenous groups had data extracted independently for each unique group. QA was conducted using the Newcastle-Ottawa Scale (NOS), adapted for observational studies [23]. To assess the risk of publication bias, the effect odds ratio (OR) for each of the included studies was plotted against their standard error on a logarithmic scale to produce a funnel plot, which were assessed for asymmetry.

### Data Analysis

A random-effects model was used to define all pooled outcome measures and the OR was estimated with its variance and 95% confidence interval (CI). The prevailing heterogeneity between ORs for comparable outcomes between different studies was calculated using the I-squared inconsistency test. The absence of statistical heterogeneity is indicated by a value of 0%, whereas larger values indicate increasing heterogeneity. Studies were only eligible for inclusion in meta-analysis if data were reported from which summary associations and their 95% CIs could be calculated. All meta-analyses were carried out using Review Manager Version 5.4 (Cochrane Collaboration, 2020).

Outcomes from studies were separated into three categories: 1) post-operative morbidity, 2) post-operative mortality, and 3) hospital stay. Morbidity included surgical and systemic infections, hematologic/thromboembolic, cardiovascular, pulmonary, genitourinary, immunologic, and procedure-specific post-operative complications (ileus, nerve injury, anesthetic complication, prosthesis-related complications, etc). Overall morbidity included all morbidities listed above pooled together, including those defined as “operative complications’’. Mortality was divided into two categories: 1) in-hospital and 30-day mortality (<30-day mortality) and 2) greater than 30-day mortality (>30-day mortality), which included mortality and survival. Overall mortality refers to both <30-day and >30-day mortality pooled together. Hospital outcomes included readmission, reoperation, and length of hospital stay (LOS).

Subgroup analyses were conducted based on surgical speciality, type of operation, geography, and quality of study. We also conducted a sub-group analysis by date of publication (before January 1st 2017 versus after January 1st 2017). This publication period sub-group analysis was done to compare studies before and after both declarations for Indigenous rights were published in North America (Truth and Reconciliation Commission (TRC), Canada, 2015; American Declaration of the Rights of Indigenous Peoples (ADRIP), USA, 2016) to see if we could detect a difference in surgical outcomes over time, specifically after increased advocacy for Indigenous people on this continent [24,25]. Sensitivity analysis compared fixed effects to random effects models to test the assumption that the random effects method was the most appropriate choice. A study could contribute to more than one analysis if it reported on multiple outcomes (i.e., overall morbidity AND surgical site infections AND mortality analyses).

## RESULTS

### Study Selection and Characteristics

A PRISMA-ScR flow diagram outlining the scoping review process is presented in Figure 1. The initial search resulted in a total of 11423 non-duplicate studies, of which 697 were included in full-text review after title and abstract review. Following full-text review, expert consultations, and relevant review appraisal for additional relevant articles, 105 unique studies met inclusion criteria. Twenty-one studies were multiple publications, meaning they reported findings on the same cohort as another study included in this review. We chose to report data only from the original study, or from the study that covered the most extensive cohort, to prevent repetition of data points, and multiple publications were not included in the meta-analysis or narrative review.

**Fig 1.**
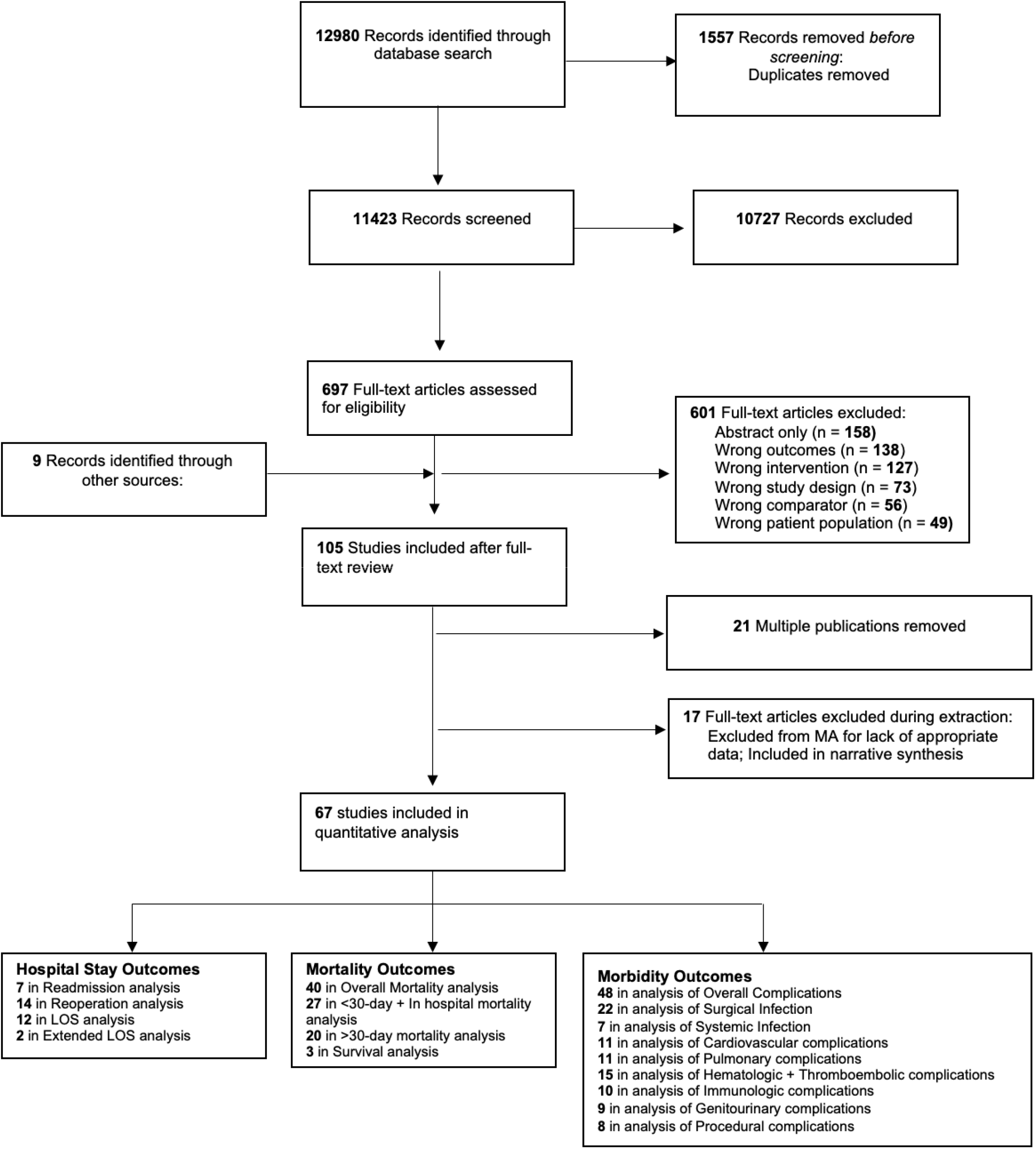
PRISMA-ScR flow diagram of study selection process, inclusions, and exclusions.

Of the 84 studies included in the narrative synthesis, 36 were retrospective cohort studies, 7 were prospective cohort, 4 were case-control, and 36 were cross-sectional. A comprehensive summary of findings and characteristics of all included studies are presented in Table 1 and S1 Table. For a complete list of references, see S3 Appendix.

**Table 1.**
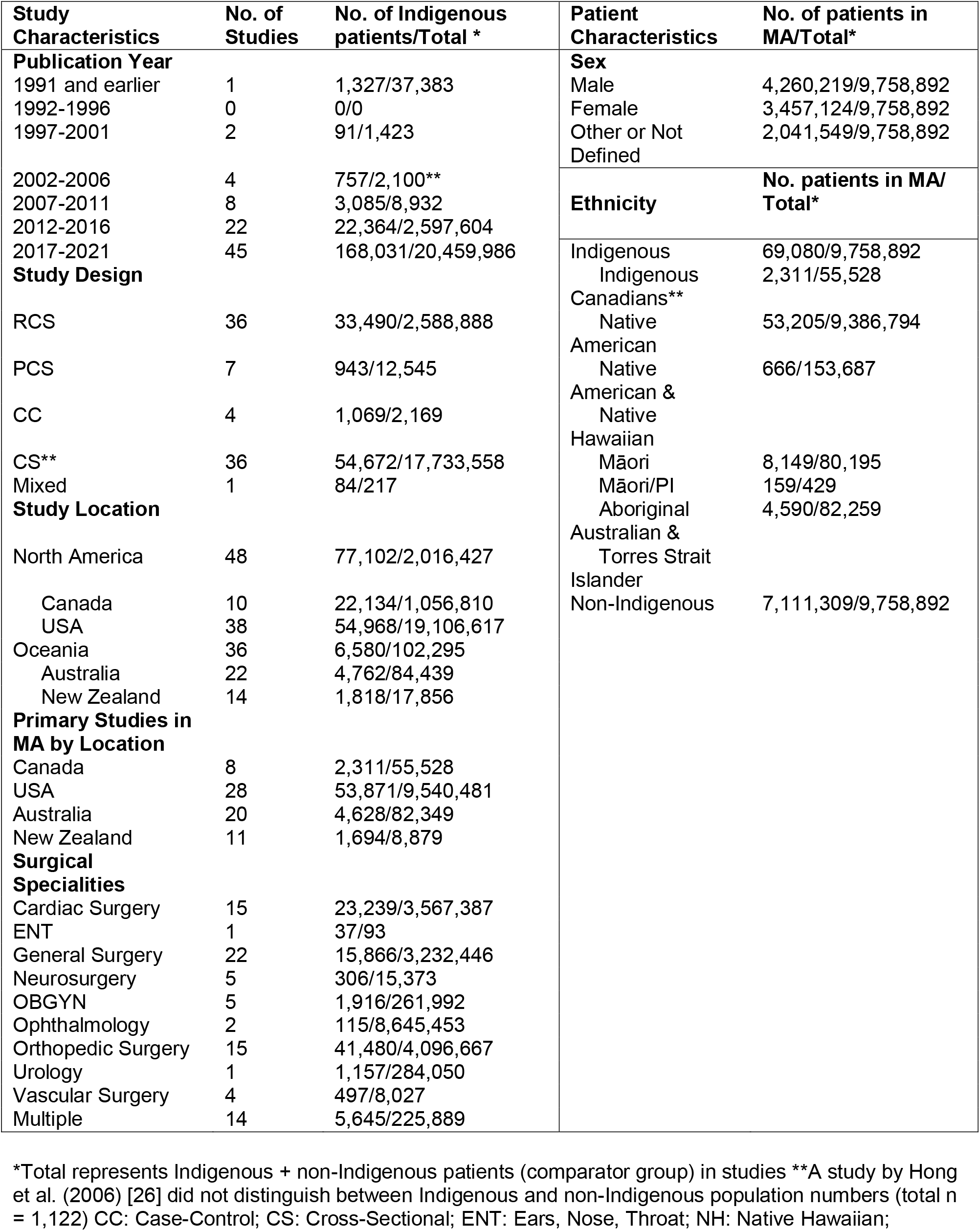
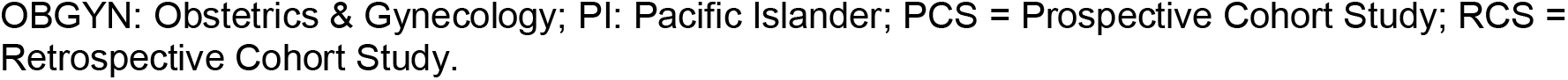
Study and patient characteristics.

Studies were published between 1989 and 2021, with research conducted from 1971 to 2019. A total of 37 studies were published over the first 28 years (1989-2016), while 45 were published in the last 5 years (2017-2021). 36/84 (42.9%) studies were based in Oceania (22/36 (61.1%) in Australia and 14/36 (38.9%) in New Zealand) while 48/84 (57.1%) studies were based in North America (38/48 (79.2%) in USA and 10/48 (20.8%) in Canada). Surgical outcomes were reported for 9,758,892 patients across 9 surgical specialties, including General Surgery (n=22 studies), Orthopedic Surgery (n=15 studies), Cardiac Surgery (n=15 studies), Urology (n=1 studies), Obstetrics and Gynecology (n=5 studies), Neurosurgery (NS) (n=5 studies), Vascular Surgery (n=4 studies), Ears, Nose, and Throat (ENT) (n=1 studies), and Ophthalmology (n=2 studies). Fourteen studies reported surgical outcomes from multiple surgical specialties. A breakdown of procedures can be found in S2 Table.

A total of 9,758,892 patients across 67 studies were included in the meta-analysis, of which 69,080 (0.7%) were Indigenous and 7,111,3091 (99.3%) were non-Indigenous. Indigenous populations consisted of Native American (n=53,205, 77.0%), Māori (n=8,194, 11.9%), Aboriginal Australians and Torres Strait Islanders (n=4,590, 6.6%), and Indigenous Canadians (n=2,311, 3.3%). Some studies grouped

Indigenous populations together, reporting on Māori and Pacific Islanders (n=159, 0.2%), or Native Hawaiian and Native American groups (n=666, 1.0%).

### Risk of Bias Assessment and Sensitivity Analyses

Half of included studies (42/84, 50%) were low quality and the other half (42/84, 50%) were good quality (S4 Appendix)). The low quality of studies was mainly attributed to failure of studies to control for confounders such as age, pre-existing comorbidities, and/or sex (40/42; 95.2%). Funnel plots for each outcome were generated, however due to the inherent heterogeneity of the studies included in each outcome category, asymmetry could not be reliably assessed (S5 Appendix). No noticeable change in the direction of the effect with a fixed effects method was appreciated, therefore a random effects model was used.

### Post-Operative Morbidity

Fifty-four studies provided data on post-operative morbidity. Of these, 48 were included in the meta-analysis and 6 were included in the narrative synthesis. Overall, there was a significantly increased morbidity for the Indigenous cohort (OR=1.26, 95% CI: 1.10-1.44, p=0.001). When low quality studies were excluded, there was 1.30 increased odds of post-operative morbidity among Indigenous compared to non-Indigenous patients (OR=1.30, 95% CI:1.12-1.51, p<0.001) (Fig 2). [29-50] When stratified by country, overall post-operative morbidity remained significantly higher in Indigenous groups from Australia (OR=1.42, 95% CI: 1.07-1.90, p=0.02) and New Zealand (OR=1.63, 95% CI: 1.09-2.43, p=0.02) compared to non-Indigenous, but there was no significant difference in overall morbidity between Indigenous and non-Indigenous patients from Canada (OR=1.71, 95% CI: 0.90-3.24, p=0.10) nor the USA (OR=1.07, 95% CI: 0.87-1.32, p=0.53).

**Fig 2.**
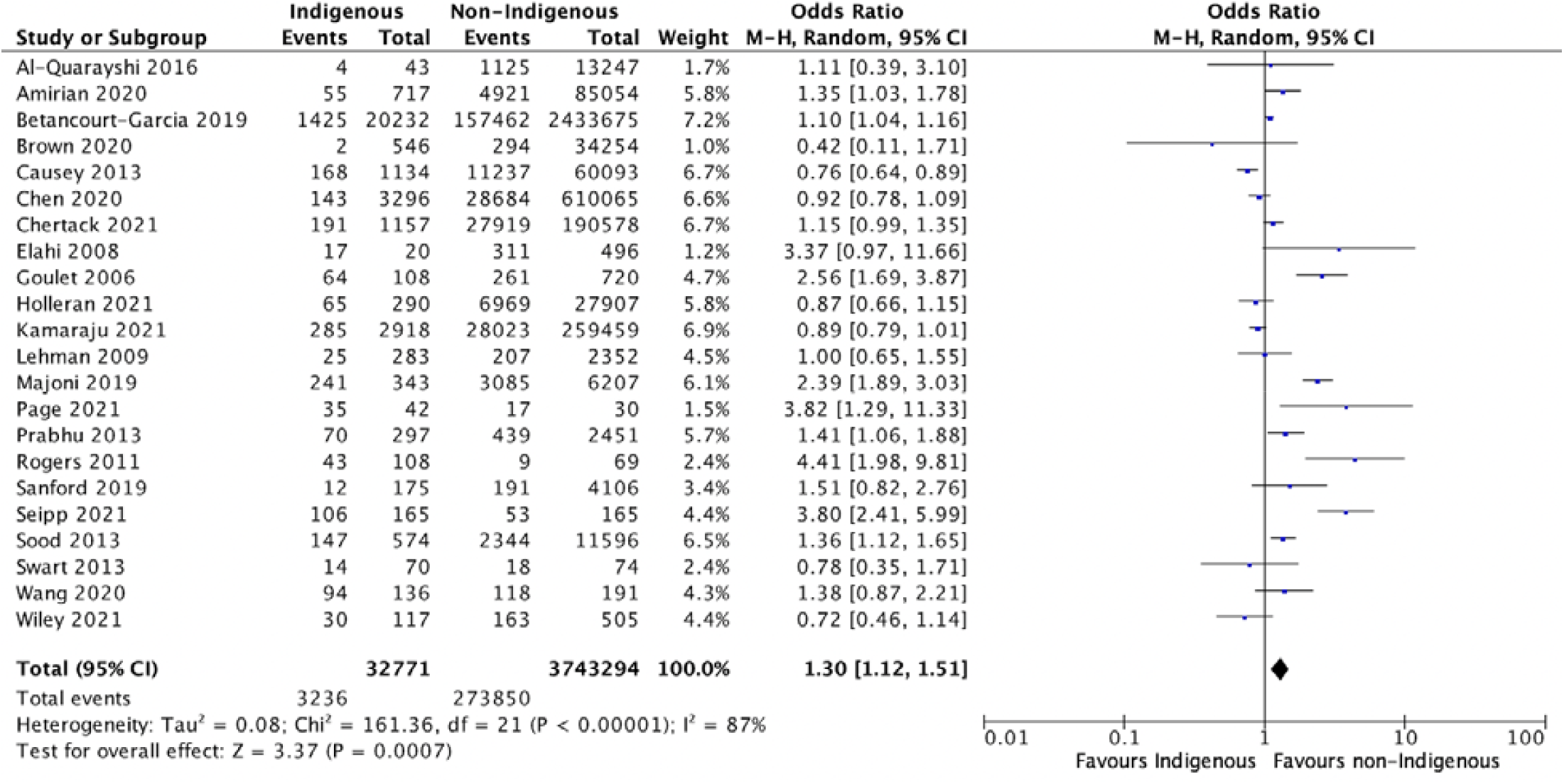
Meta-analysis of overall morbidity with good quality studies. Results are depicted for meta-analysis using a random-effects model for odds of overall morbidity post-operatively between Indigenous and non-Indigenous surgical patients. Overall morbidity included any post-operative complication including surgical infection, systemic infection, cardiovascular complication, pulmonary complication, hematologic/thromboembolic complication, genitourinary complication, immunologic complication, and/or procedural complication. This analysis was conducted using good quality observational studies, as determined by Newcastle Ottawa Scale ratings.

When separated by surgical specialty, urology (OR=1.79, 95% CI:1.32-2.41, p<0.001) and cardiovascular surgery (OR=1.42, 95% CI: 1.11-1.82, p=0.005) had increased overall morbidity in Indigenous patients.

Specifically, renal transplants (OR=1.97, 95% CI:1.34-2.89, p<0.001) and coronary artery bypass grafts (OR=1.50, 95% CI: 1.27-1.76, p <0.001) both resulted in significantly higher post-operative complications for Indigenous patients. No significant differences in overall morbidity were found in other surgical specialties or procedures. All results for overall morbidity and mortality meta-analyses can be found in Table 2.

**Table 2.**
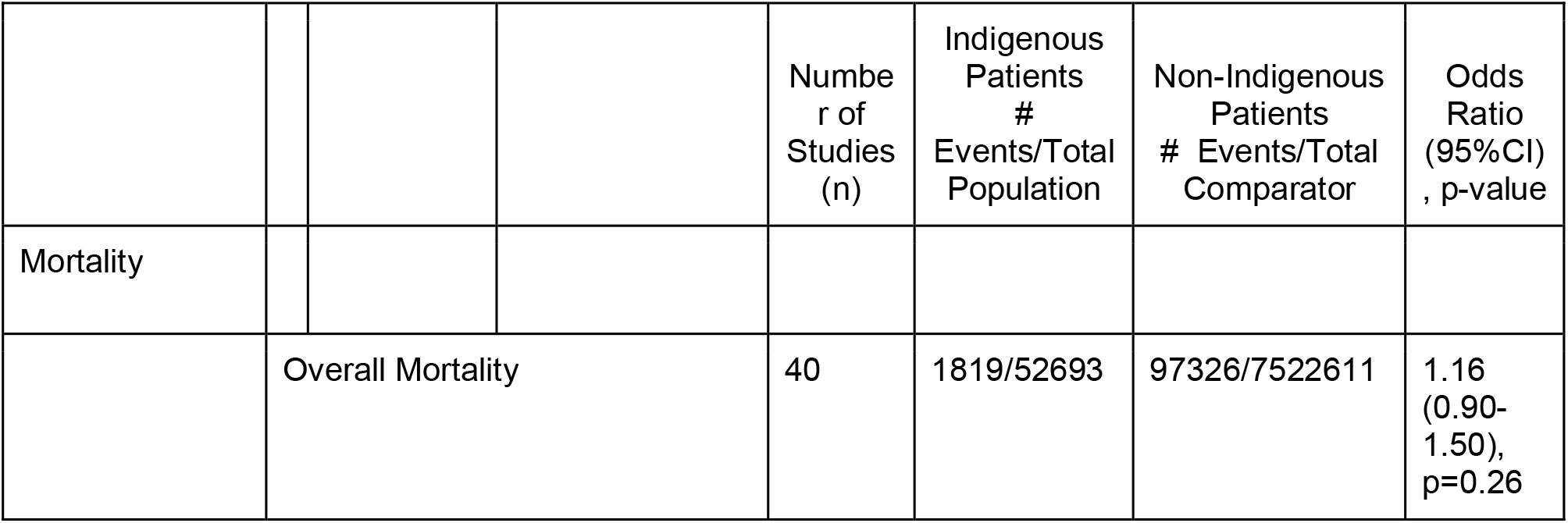
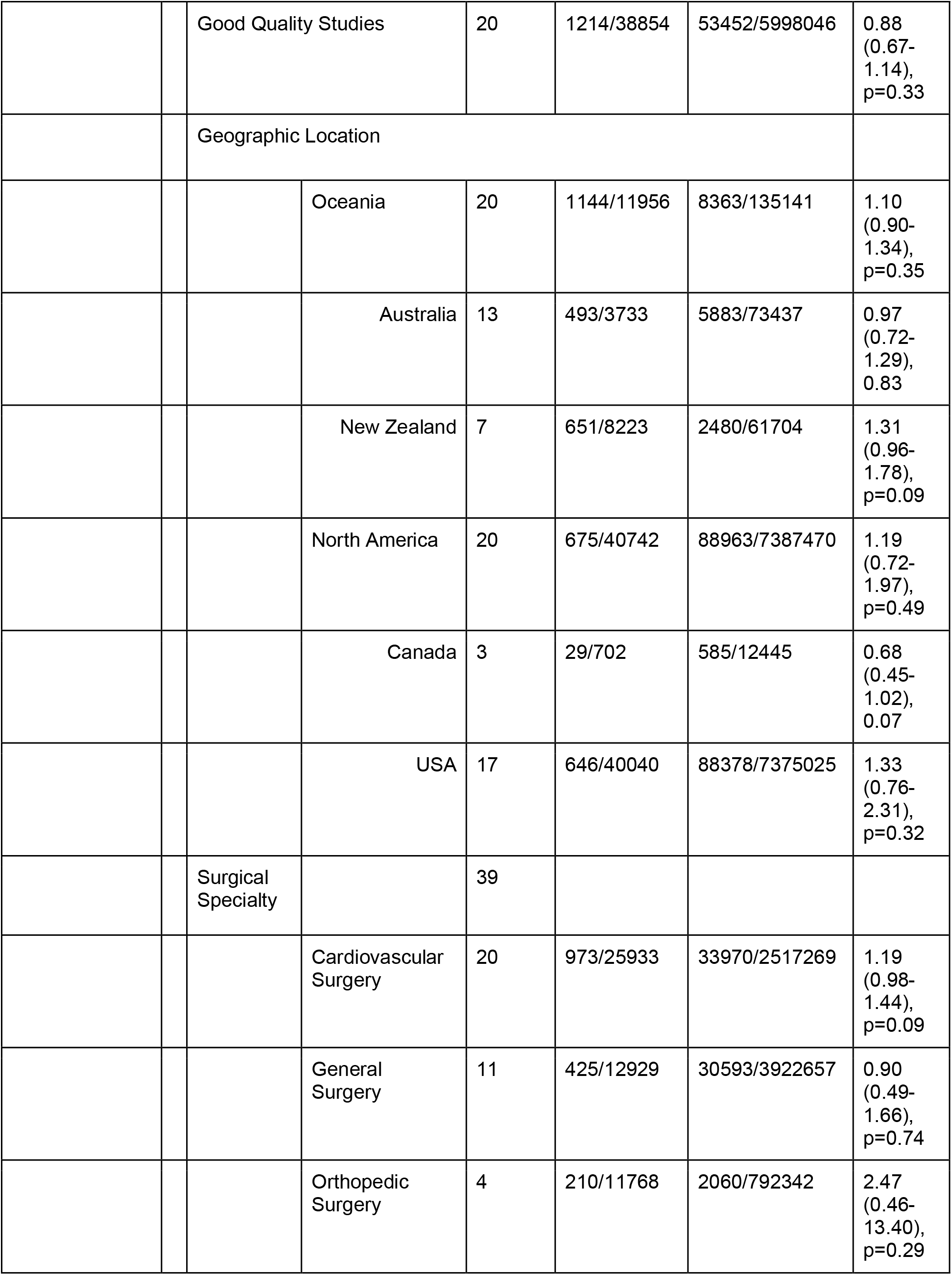
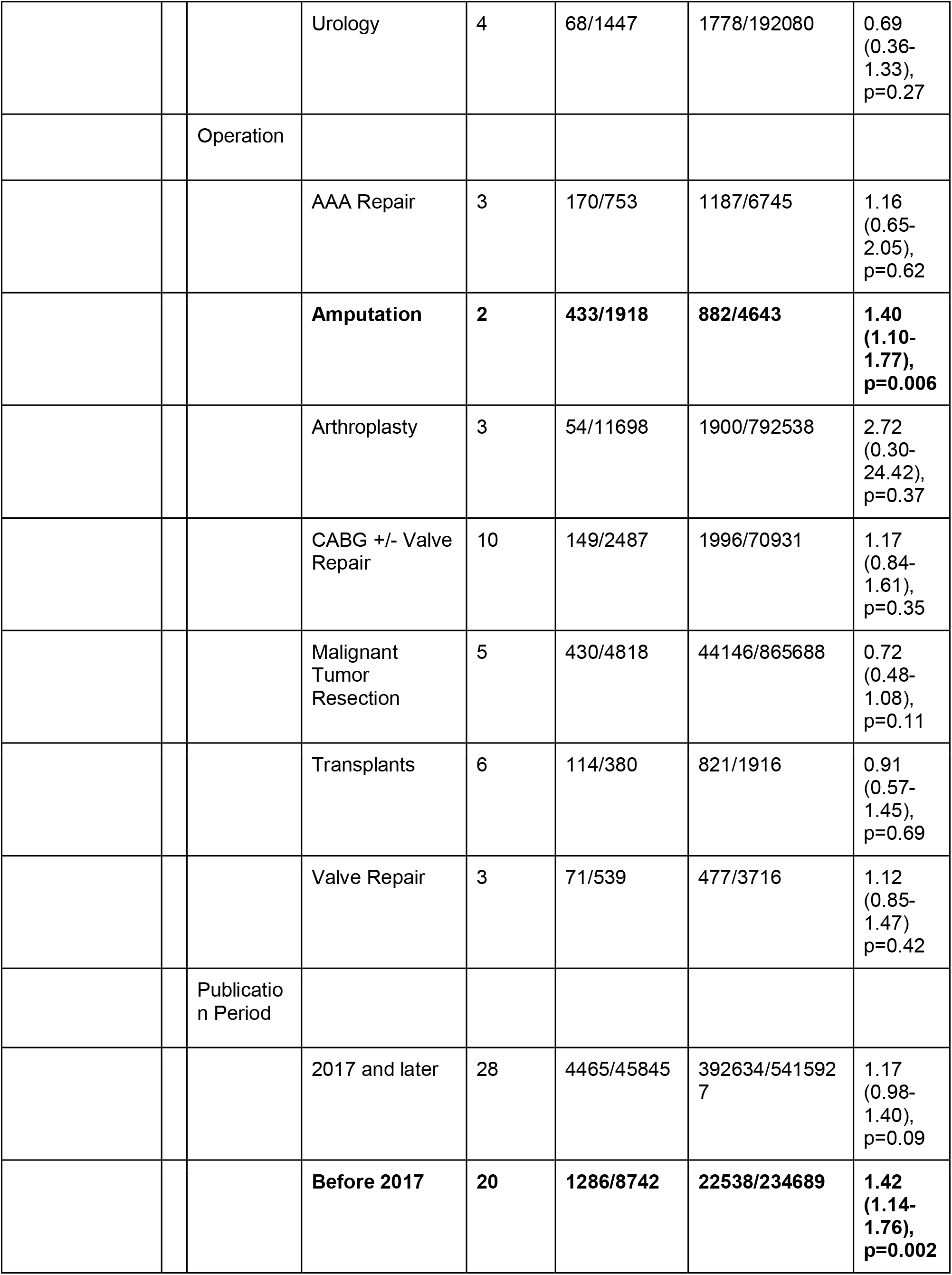
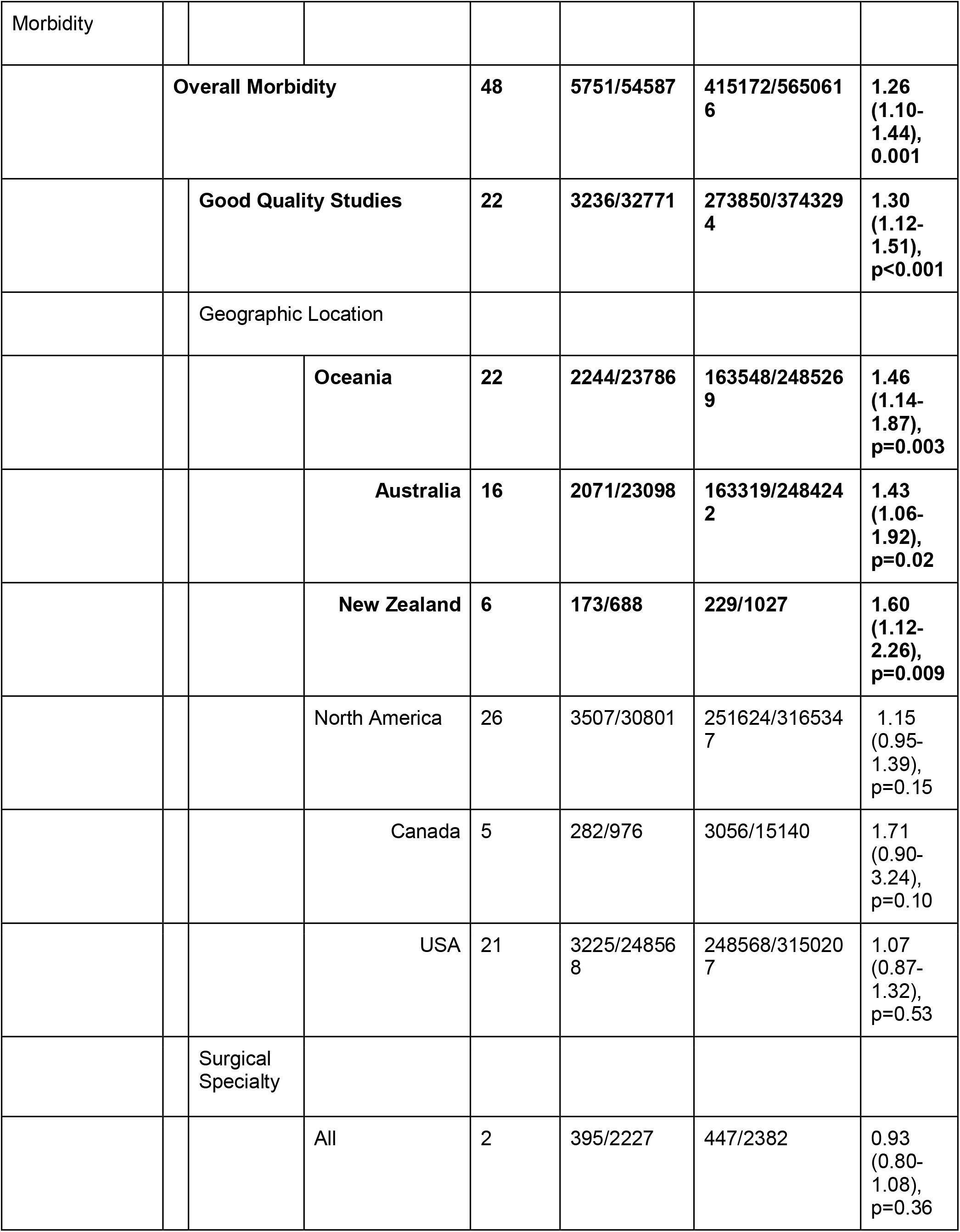
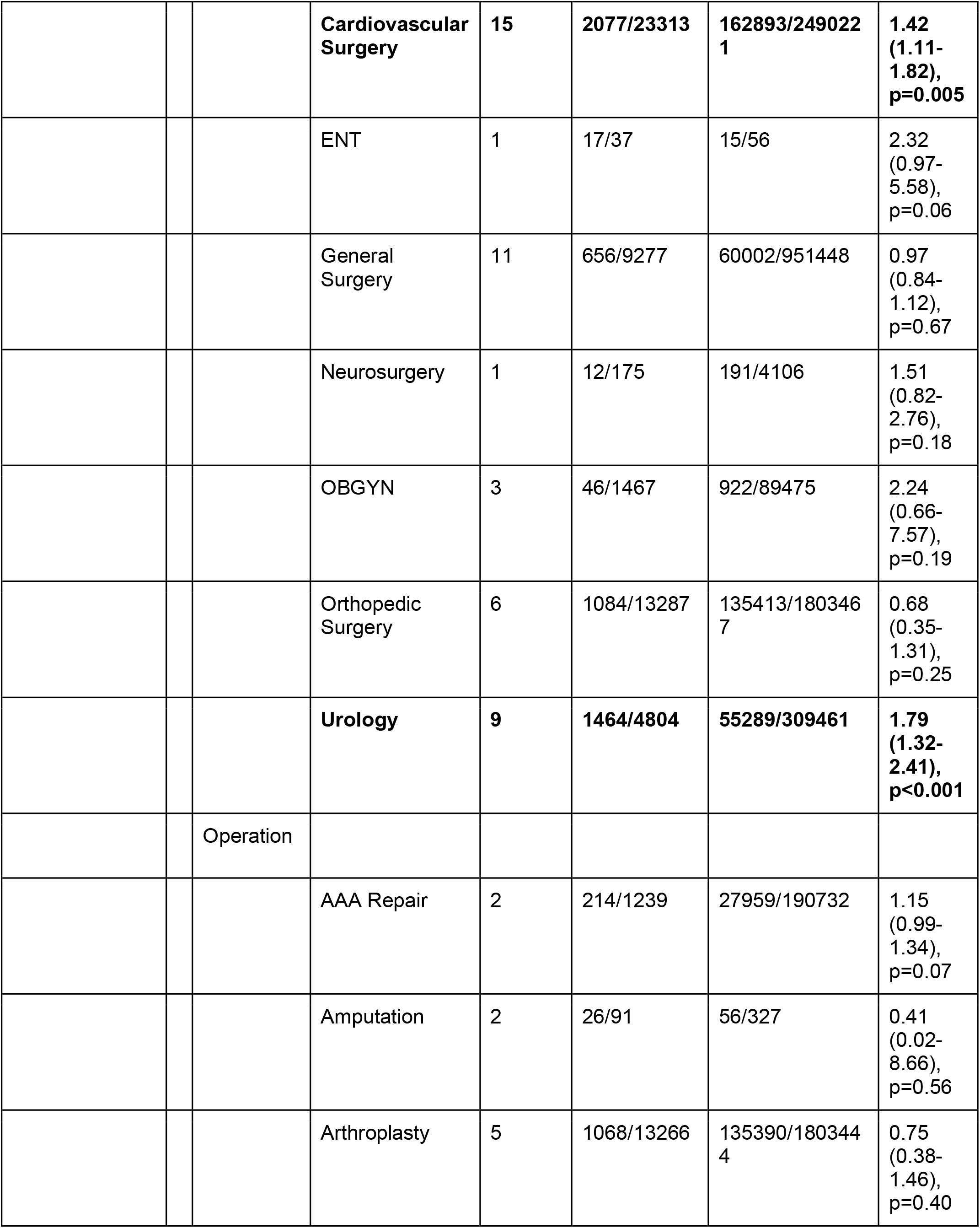
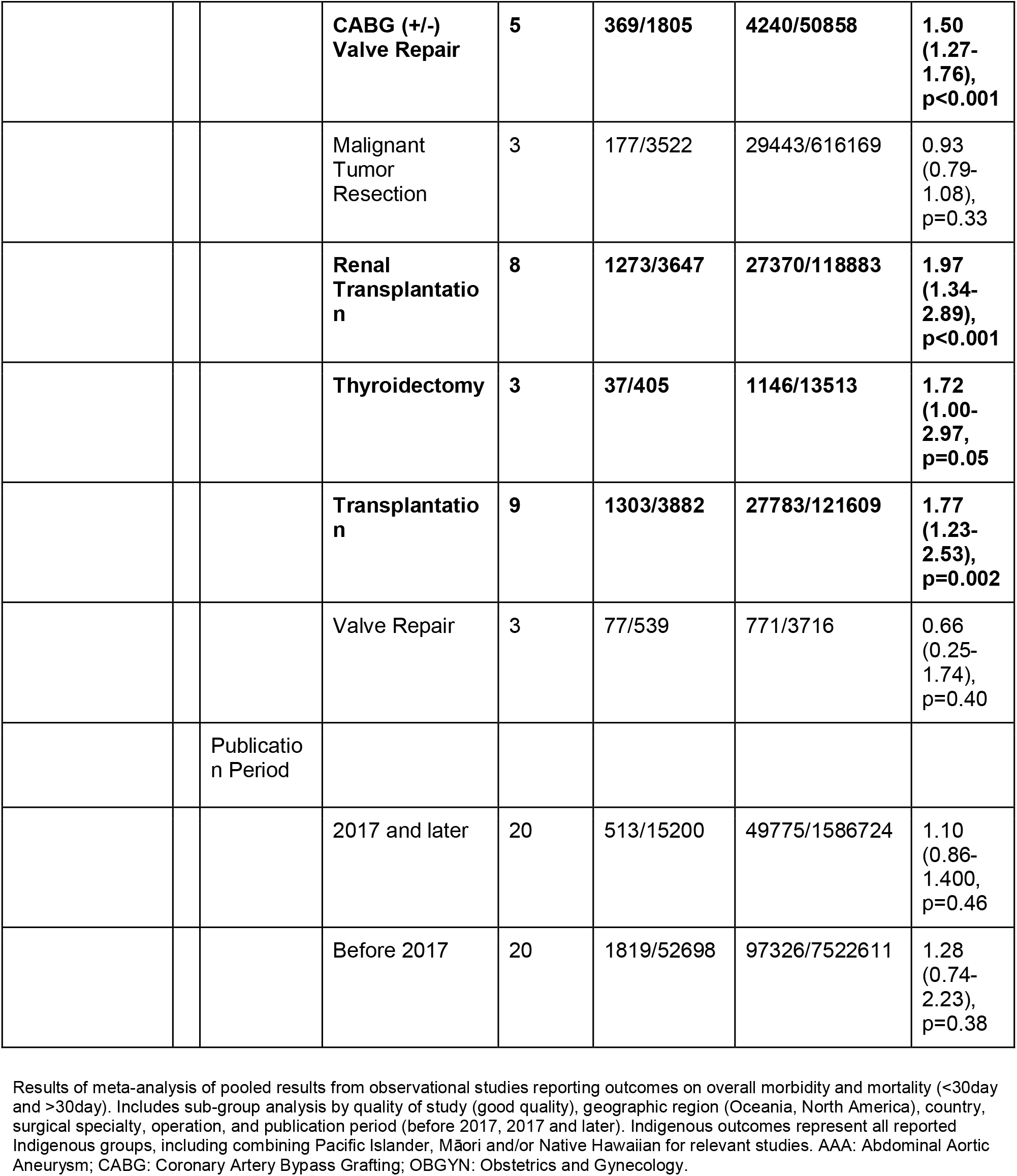
Meta-Analysis for overall morbidity and mortality outcomes.

Twenty studies reported morbidity outcomes prior to 2017 and 28 reported morbidity outcomes between January 2017 and December 2021. Before 2017, there was significantly more overall morbidity among Indigenous patients (OR=1.42, 95% CI: 1.14-1.76, p=0.002); after 2017, on the other hand, there was no significant difference in overall morbidity (OR=1.17, 95% CI: 0.98-1.40, p=0.09). All other meta-analyses results are in S3 Table.

Of the six studies reporting morbidity outcomes from narrative analysis, four studies reported no significant differences between Indigenous and non-Indigenous patients [51-54]. The other two found significantly increased functional deficits and infection rates for the Indigenous populations [55, 56].

### Surgical and Systemic Infections

Surgical site and systemic infection data were included in 29 unique studies. Indigenous patients had 1.34 times increased odds of post-surgical infections compared to non-Indigenous patients (OR=1.34, 95% CI: 1.12-2.59, p=0.001). When stratified by continent, North American Indigenous Peoples had 1.33 times increased odds of post-surgical infections compared to non-Indigenous North Americans (OR=1.33, 95% CI: 1.10-1.60, p=0.003) (Figure 3a) [30,31,36,39,40,42-44,47,48,57]. When stratified by country, only Indigenous Peoples from the USA experienced significant disparities in surgical infection rates (OR=1.36, 95% CI: 1.06-1.74, p=0.01). There were no differences in post-operative systemic infections detected in the analyses. There were no significant differences in post-operative surgical site infections (OR=1.16, 95% CI: 0.90-1.49, p=0.24) or systemic infections (OR=1.01, 95% CI: 0.58-1.75, p=0.98) in studies published before 2017. In studies published after 2017, there was a significant increase in surgical site infections for Indigenous patients (OR=1.5, 95% CI: 1.16-1.95, p=0.002), but not in systemic infections (OR=1.01, 95% CI:0.92-1.10, p=0.88).

**Fig 3.**
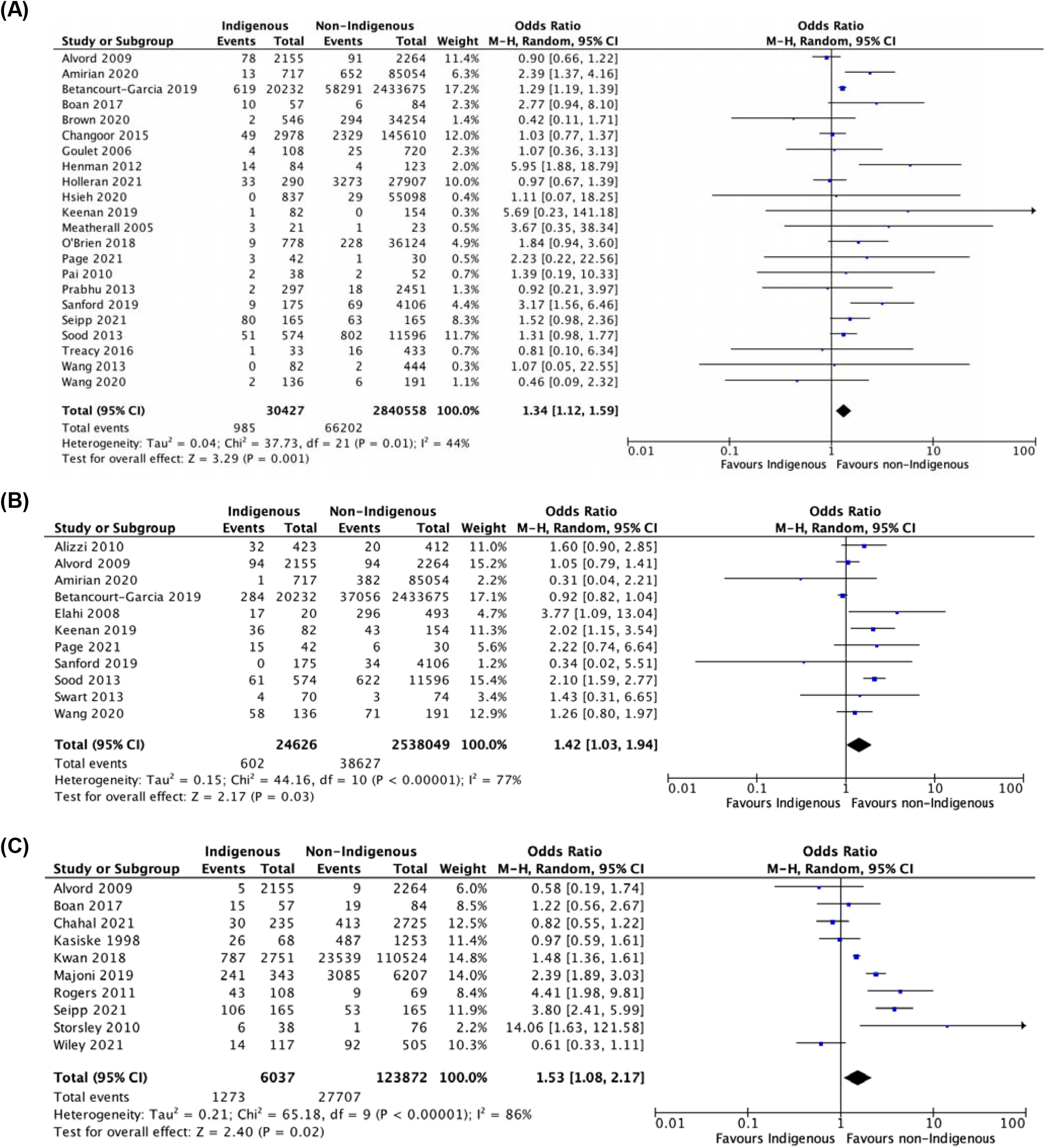
Meta-analyses with studies for post-operative (A) Surgical infections; (B) Pulmonary complications; and (C) Immunologic complications. Results are depicted for meta-analysis using a random-effects model for observational studies. A. post-operative surgical infections including superficial, organ space, and deep wound infections, wound dehiscence, abscess formation; B. pulmonary complications including pneumonia, respiratory failure, and prolonged ventilation and C. immunologic complications including graft failure, delayed graft function, and graft rejection (acute, chronic).

### Other Post-Operative Complications

Indigenous Peoples had higher odds of pulmonary complications if all studies were included (OR=1.42, 95% CI: 1.03-1.94; p=0.03, Fig 3b). If only good quality studies were included, however, this difference was not significant (OR=1.32, 95% CI: 0.92-1.90, p=0.13). Patients undergoing cardiovascular surgery (OR=1.85, 95% CI:1.50-2.29, p<0.001) and Oceanic Indigenous patients undergoing any surgery (OR=1.64, 95% CI:1.25-2.17, p<0.001) had higher odds of pulmonary complications compared to non-Indigenous peoples. Furthermore, there was a significant increase in pulmonary complications in Indigenous patients in studies published prior to 2017 (OR=1.64, 95% CI:1.06-2.54, p=0.02), while this was not significant in studies published in the last five years (OR=1.22, 95% CI:0.81-1.83, p=0.34). There were no geographic or surgical specialty differences between populations with respect to cardiovascular complications.

Indigenous patients were significantly more likely to experience immunologic complications post-operatively (OR=1.53, 95% CI:1.08-2.17, p=0.02) (Fig 3c) [35,36,40,42,46]. Oceanic Indigenous Peoples (OR=2.35, 95% CI: 1.36-4.04, p=0.002) and those undergoing renal transplantation (OR=1.55, 95% CI: 1.44-1.67, p<0.001) had higher odds of these complications. Furthermore, studies published in the last five years demonstrated a higher rate of immunologic complications for Indigenous patients (OR=1.48, 95% CI:1.00-2.20), p=0.05) while this was not significant in studies published before 2017 (OR=1.96, 95% CI:0.64-6.00, p=0.24). There were no significant differences in genitourinary (OR=1.21, 95% CI: 0.92-1.60, p=0.18), hematologic (OR=1.28, 95% CI:0.97-1.70, p=0.08), or procedural complications (OR=0.96, 95% CI: 0.32-2.92, p=0.94) between Indigenous and non-Indigenous patients. Further subgroup analyses by geographic region, surgical specialty, quality of study, and publication period likewise did not demonstrate significant difference in these outcomes (S3 Table).

### Post-Operative Mortality and Survival

Fifty studies provided information on post-operative mortality and/or survival. Of these studies, 40 were included in the meta-analysis, totalling 52,698 Indigenous and 5,057,266 non-Indigenous post-operative deaths. Overall mortality was similar for both Indigenous and non-Indigenous patients (OR=1.16, 95% CI: 0.90-1.50, p=0.26), and there were no significant differences between the groups based on geography, surgical specialty, or quality of study (Table 2). Results of analysis on <30-day mortality (OR=1.20, 95% CI: 0.81-1.78, p=0.37) and >30-day mortality (OR=1.16, 95% CI: 0.95-1.41, p=0.15) demonstrated similar mortality rates for Indigenous and non-Indigenous patients. However, Oceanic patients (OR=1.29, 95% CI: 1.06-1.57, p=0.01), and more specifically, Māori and PI patients from New Zealand (OR=1.39, 95% CI: 1.01-1.92, p=0.04) had higher odds of >30-day mortality than non-Indigenous patients. Indigenous patients undergoing orthopedic surgeries and, more specifically, amputations, had increased odds of <30-day mortality versus non-Indigenous patients (OR=1.32, 95% CI: 1.08-1.61, p=0.006 and OR=1.40, 95% CI: 1.10-1.77, p=0.006). Of the 10 studies included in narrative analysis, only one found increased mortality rates for Indigenous patients (HR=1.15, 95% CI: 1.05-1.26, p= NR) [58]. There were no significant differences in overall mortality in studies published before (OR=1.28, 95% CI: 0.74-2.23, p=0.38) or after 2017 (OR=1.10, 95% CI: 0.86-1.40, p=0.46).

### Hospital Stay Outcomes

Forty-five studies were included in the meta-analysis for hospital stay outcomes. There was a significant increase in reoperation rates (OR=1.33, 95% CI: 1.02-1.74, p=0.04) and LOS for Indigenous patients (SMD=0.15, 95% CI: 0.02-0.29, p=0.02). The difference in reoperation rates (OR=1.33, 95% CI: 1.02-1.74, p=0.03) and LOS (SMD=0.65, 95% CI: 0.14-1.16, p=0.01) before 2017 was statistically significant, while the difference in reoperation rates (OR=1.35, 95% CI: 0.61-2.97, p=0.46) and LOS (SMD=0.02, 95% CI: -0.04-0.08, p=0.53) after 2017 was not statistically significant. Oceanic Indigenous patients had the highest odds of reoperation and longest LOS (SMD=0.54, 95% CI -0.00-1.08, p=0.05). There were no significant differences in readmission rates for either population group (S3 Table). The three studies included in narrative analysis did not report significant differences in hospital stay outcomes for Indigenous patients [51,53,60].

## DISCUSSION

This study presents a comprehensive overview and summary of the state of post-operative outcomes for Indigenous populations worldwide. Our findings are consistent with existing literature that describes disparate post-operative outcomes for Indigenous patients [14,16]. Specifically, this study presents evidence that Indigenous patients in Canada, the USA, Australia, and New Zealand experience greater post-operative morbidity, including infections and other systemic complications, than their non-Indigenous counterparts. We also found that long-term mortality was significantly increased for Indigenous patients from New Zealand, echoing existing literature [61,62]. Additionally, this study highlighted a high proportion of low-quality studies on the topic, as well as a very low representation of Indigenous patients in published research, which is a call to action for researchers to scrutinize this topic more thoroughly.

Specifically, this study found that Indigenous people in Australia and New Zealand had significantly higher post-operative morbidity and mortality compared to non-Indigenous peoples. These differences were not statistically significant in Canadian and American populations. However, previous studies, with differing inclusion and exclusion criteria, have highlighted disparities between Indigenous and non-Indigenous surgical outcomes in Canada and the USA [16,65].

On an international stage, the UNDRIP was adopted by the UN General Assembly in 2007 to protect Indigenous Peoples worldwide, including enshrining the right of Indigenous Peoples “without discrimination, to … health and social security” [13]. As previously mentioned, Canada, the USA, Australia, and New Zealand were the only four countries to vote against this Declaration in 2007 [13]. However, all four countries have since ratified and given their support for the Declaration, as well as adopted their own national frameworks [24,25]. On a national level in North America, the TRC and ADRIP called on the healthcare sector to recognize, measure, and close the gaps in Indigenous health outcomes [24,25]. To date, these calls to actions have not meaningfully improved the reporting of Indigenous health outcomes in the North American context, as evidenced by the preponderance of low-quality studies retrieved in this study and the trend of increased post-operative infections and immunological complications in Indigenous patients in the last five years. However, other metrics of post-operative morbidity have improved in the last five years, such as non-significant differences in overall morbidity, hospital LOS, and reoperation rates for Indigenous patients. These conflicting results illustrate that it remains to be seen if the TRC, UNDRIP, and/or ADRIP will lead to the structural changes needed in healthcare systems to ameliorate surgical inequities for Indigenous patients.

On a more positive note, there demonstrably has been more interest in studying these inequities, as evidenced by the exponential rise in publications centered on this topic and inclusion of Indigenous patients in studies since these legislations have been adopted. In our study, we found that the publication rate doubled over the last decade, from 22 in 2012-2016 to 45 in 2017-2021, and that this translated into a seven-fold increase in the Indigenous population studied (22,364 vs 168,031 patients). In the USA, many of these publications were made possible thanks to the harnessing of data and statistical power from the National Surgical Quality Improvement Program (NSQIP), created by the American College of Surgeons (ACS) [66]. However, despite Canadian institutions having access to NSQIP since as early as 2011, none of the Canadian studies utilized NSQIP, which may partially explain the poor quality of studies from Canada (n=6/9, 66.7%) compared to the USA (n=15/39, 38.5%). One possible explanation for the lack of NSQIP utilization in the Canadian context may be that the Canadian NSQIP databases do not record ethnicity/race data, while this information is recorded in USA databases. This demonstrates a lack of appropriate tools for measuring health equity, despite call to action 19 from the TRC of Canada that states “we call upon the federal government, in consultation with Aboriginal peoples, to establish measurable goals to identify and close the gaps in health outcomes between Aboriginal and non-Aboriginal communities, and to publish annual progress reports and assess long term trends” [24].

Collecting and recording Indigenous status in national databases would directly help address this call to action, and we recommend re-visiting current Canadian policies against collecting ethnicity data in order to better analyze differences in healthcare outcomes between various ethnic groups.

Systemic racism and discrimination are prevalent in post-colonial health systems. These factors likely contribute to the observed disparities in surgical outcomes in this study, rather than Indigenous culture itself [67,68]. Institutions, including healthcare systems, created by settlers were designed to benefit the colonizers and disadvantage original inhabitants. Indigenous Peoples around the world were purposefully exposed to infectious diseases, denied treatments, experienced forced sterilization, and were banned from practicing their traditional ways of healing [69,70]. We must acknowledge the ongoing effects that colonization contributes to the current and lasting socioeconomic marginalization and consequent health care disparities experienced by Indigenous populations globally [71,72]. In this way, we can confront and begin to close these gaps. As a global surgical community, it is essential to reimagine models of surgical care that confront the impacts of colonialism on underserved populations. Reimagining surgical models to comprehensively integrate contextualized, longitudinal, and community-centric methods that meet the unique needs of Indigenous patients may help to diminish these gross surgical disparities and improve the health of Indigenous patients.

## Limitations

The heterogeneity among surgical specialties, geographies, and distinct Indigenous groups challenged this meta-analysis. While we limited our search to Canada, New Zealand, Australia, and the USA, it is important to recognize the unique and varied Indigenous groups across these countries who, while facing similar health and social inequities, have distinct cultural, social, and political ways and practices. The nuances created by heterogeneity in our sample cannot be fully addressed in the scope of this article, nor is there available data to allow for sub-analyses by distinct Indigenous groups. Further, there are substantial differences in the healthcare systems in the four included countries, which undoubtedly influence health outcomes for marginalized groups. Additionally, we used “number of events” rather than “number of patients” as our population number in the morbidity analysis. Therefore, this study may have overestimated the independence of each event. A single patient may experience multiple different complications, but these were included in the analysis as independent events per patient, not number of patients who experienced complications. The results of this study are also limited by the poor quality of studies available in the literature. Indeed, half of the included studies were judged to be of poor quality, with a majority being predominantly retrospective in nature and many reporting unadjusted results.

Finally, the lived experiences of diverse Indigenous patients across the world and the true impact of racism and system inequities in healthcare the outcomes experienced by Indigenous patients cannot fully be addressed in a meta-analysis of post-surgical outcomes. While out of this study’s scope, it is important to consider the adverse impacts of systemic racism on health status prior to patients undergoing surgery, including but not limited to disparities in access to culturally appropriate primary and preventative care as well as lack of timely access to surgical care.

## CONCLUSION

This study provides evidence that Indigenous patients in Canada, New Zealand, Australia, and the USA continue to experience worse post-operative outcomes compared to non-Indigenous patients.

Furthermore, despite multiple calls to action on both national and international levels to assess and address the impacts of colonialism and health inequities for Indigenous populations, the availability of good quality studies on surgical health of Indigenous patients is limited. In order to address the inequities in post-operative outcomes for Indigenous Peoples, we must re-imagine models of surgical care that comprehensively integrate preventative and long-term post-operative care and prioritize accessible, feasible, and culturally appropriate care for Indigenous groups. Lastly, while there has been a significant increase in studies focused on and including Indigenous patients since 2017, further research is needed to investigate the upstream adverse impacts of systemic racism on Indigenous health prior to patients undergoing surgery.

## Data Availability

All relevant data are within the manuscript and its supporting information files.

## ACKNOWLEDGMENTS

The authors respectfully acknowledge that this review was conducted on Treaty 6 territory, the traditional land of the Nêhiyawak (Cree), Anishinaabe (Saulteaux), Niitsitapi (Blackfoot), Métis, Dene, and Nakota Sioux Peoples, the unceded territories of the x□məθk□əy□əm(Musqueam), S□wx□wú7mesh (Squamish), səlilwətaD (Tsleil-Waututh), and Tsimshian Nations, traditional territory of the Lheidli T’enneh, part of the Dakelh (Carrier) First Nation, as well as the traditional lands of the Sinixt, the Ktunaxa, the Secwepmec and the Syilxsince in the country now known as Canada since time immemorial.

