## Supplementary figures and images for "Post-operative morbidity and mortality in Indigenous Peoples: A scoping review and meta-analysis"

### S1 Appendix

**S1 Appendix: PRISMA-SCr and MOOSE Checklists**

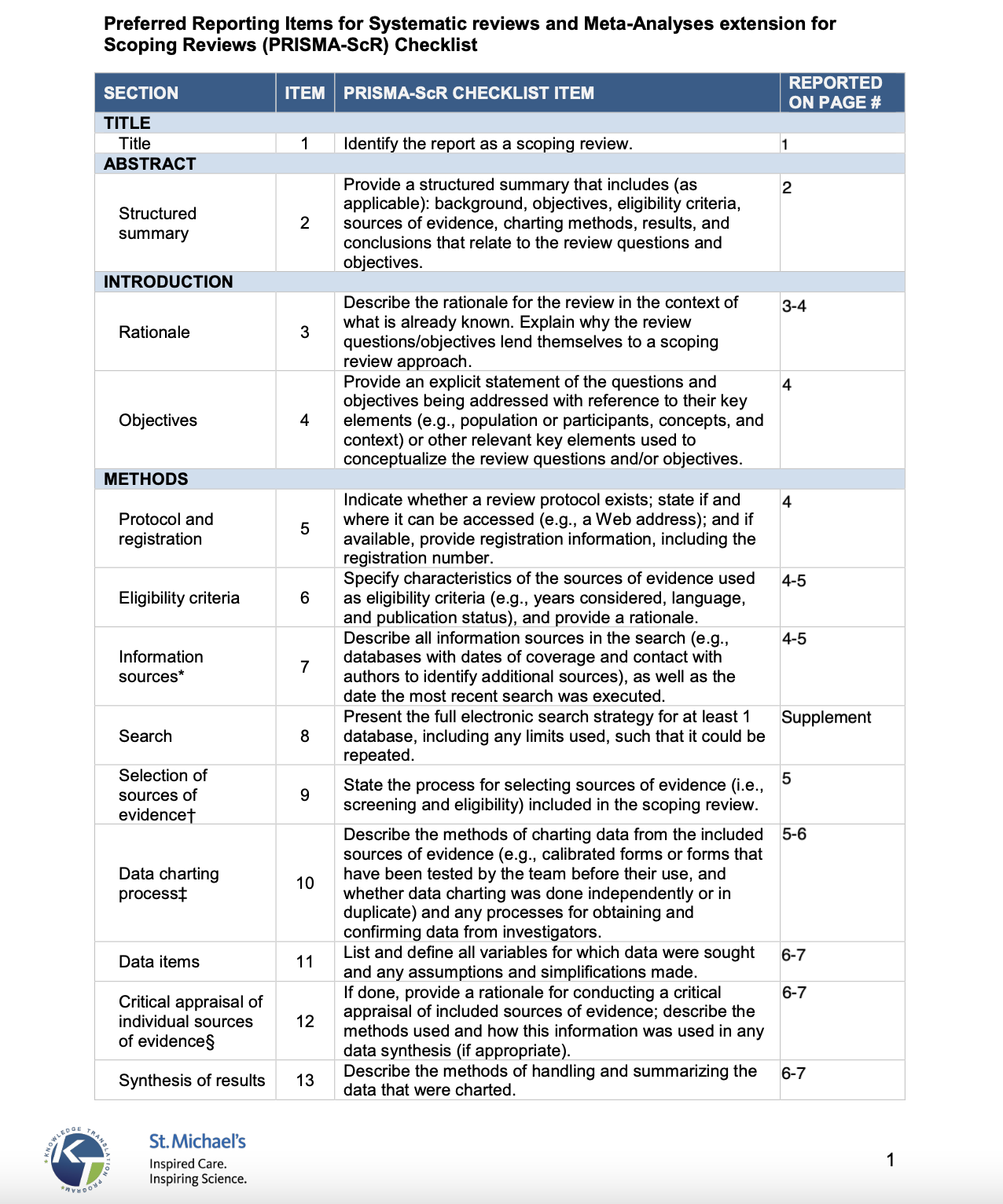

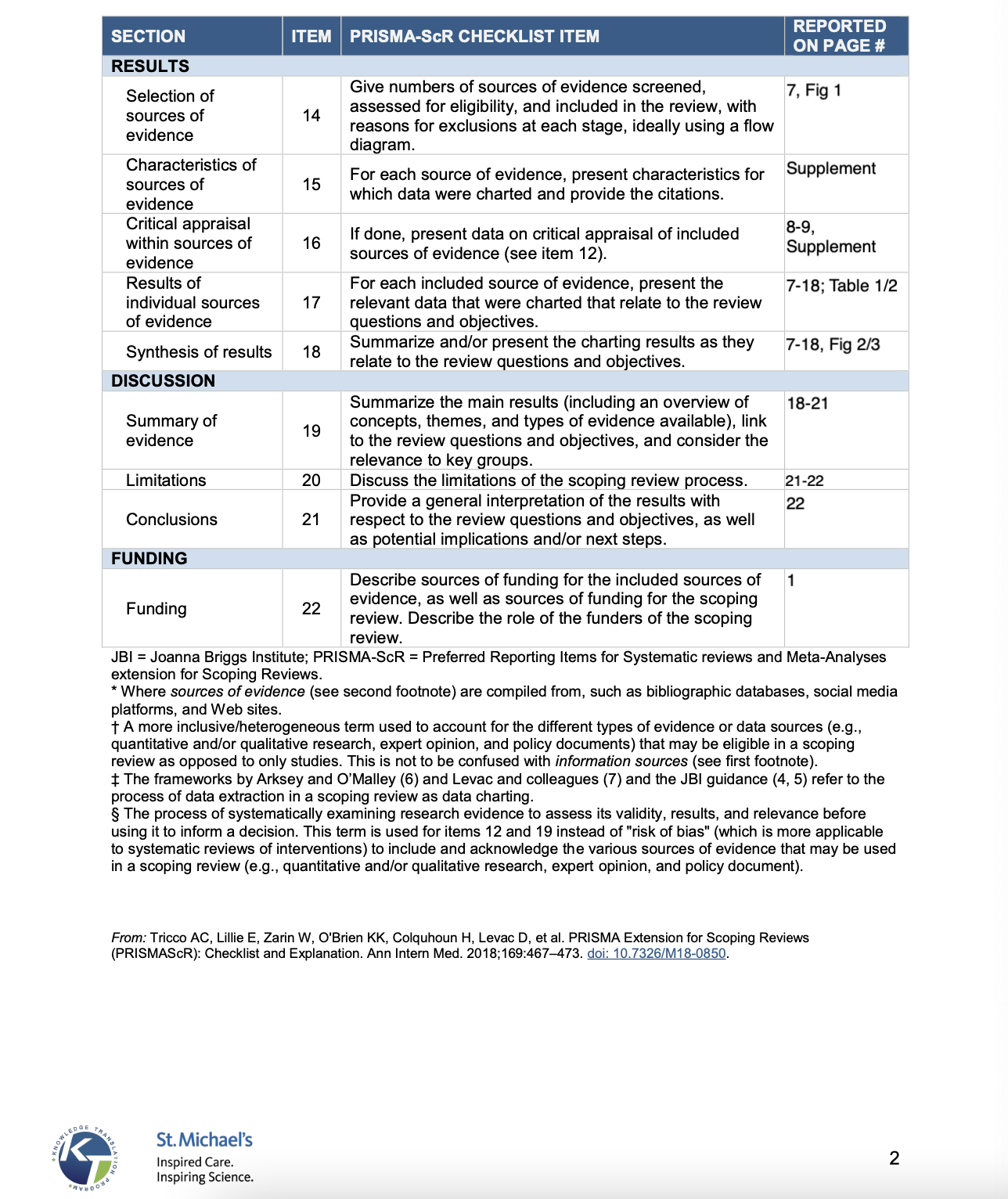

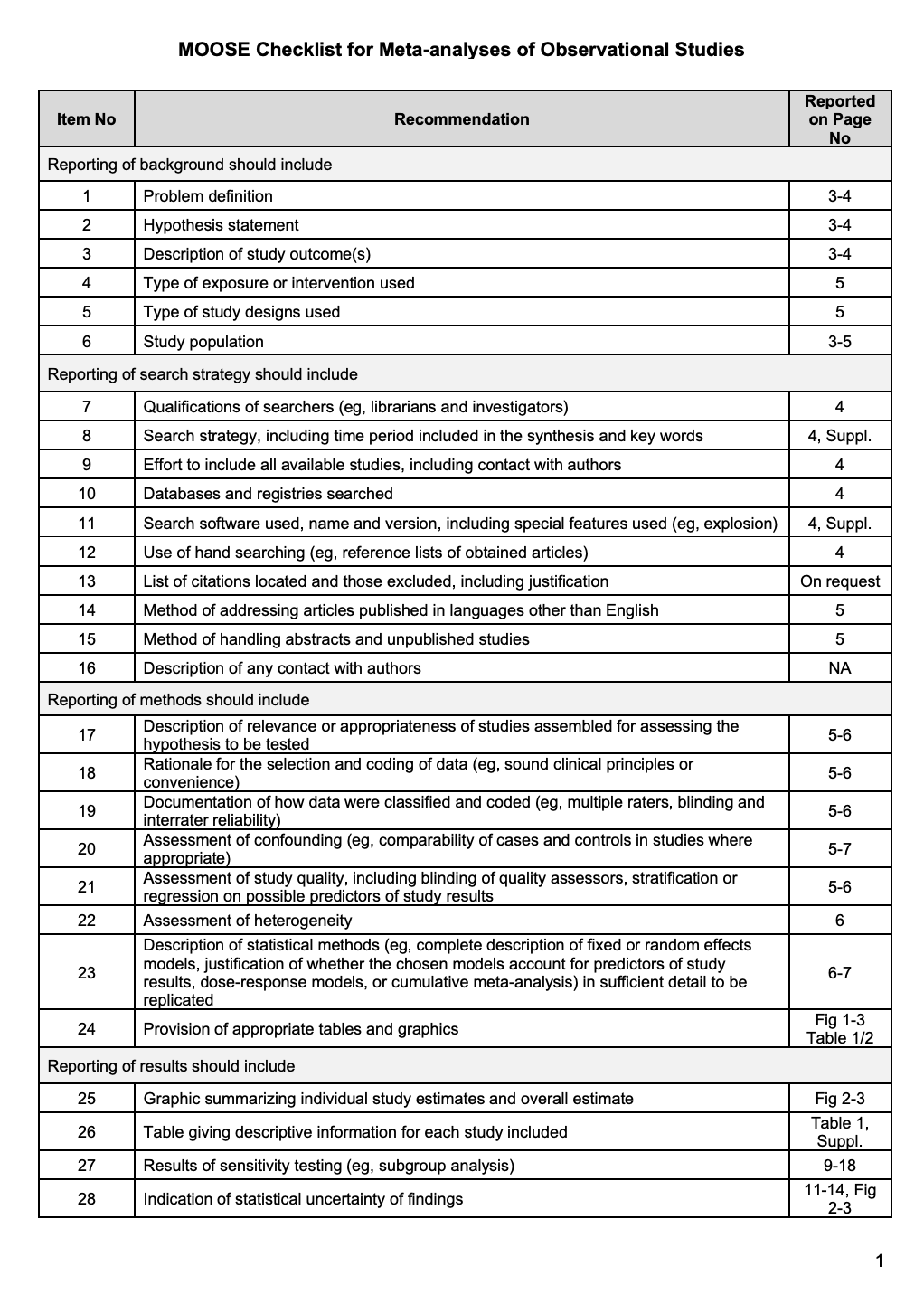

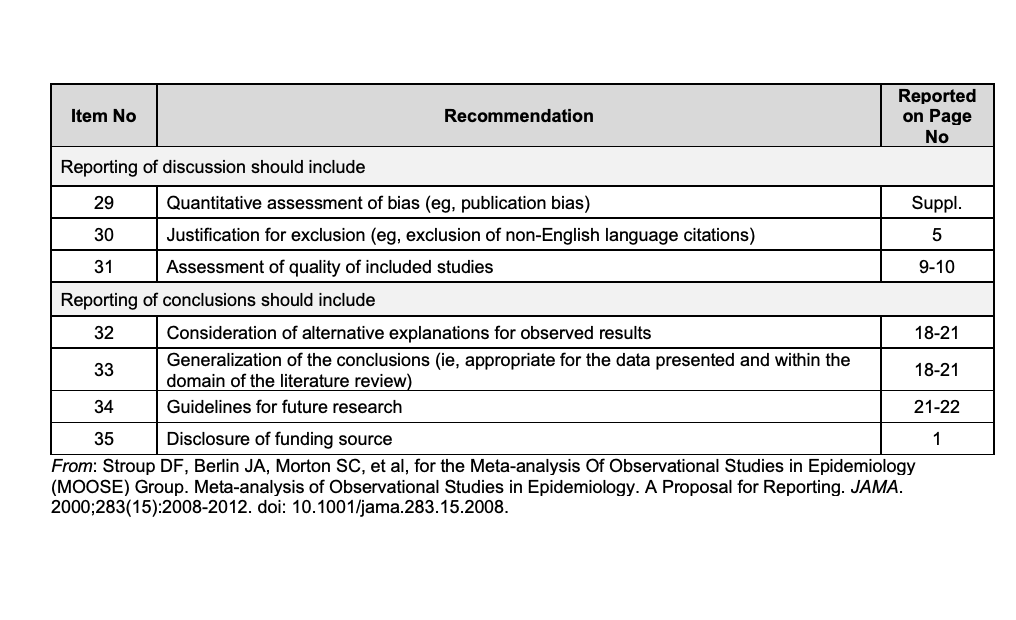

### S4 Appendix

**S4 Appendix: Quality and risk of bias assessment of included studies**

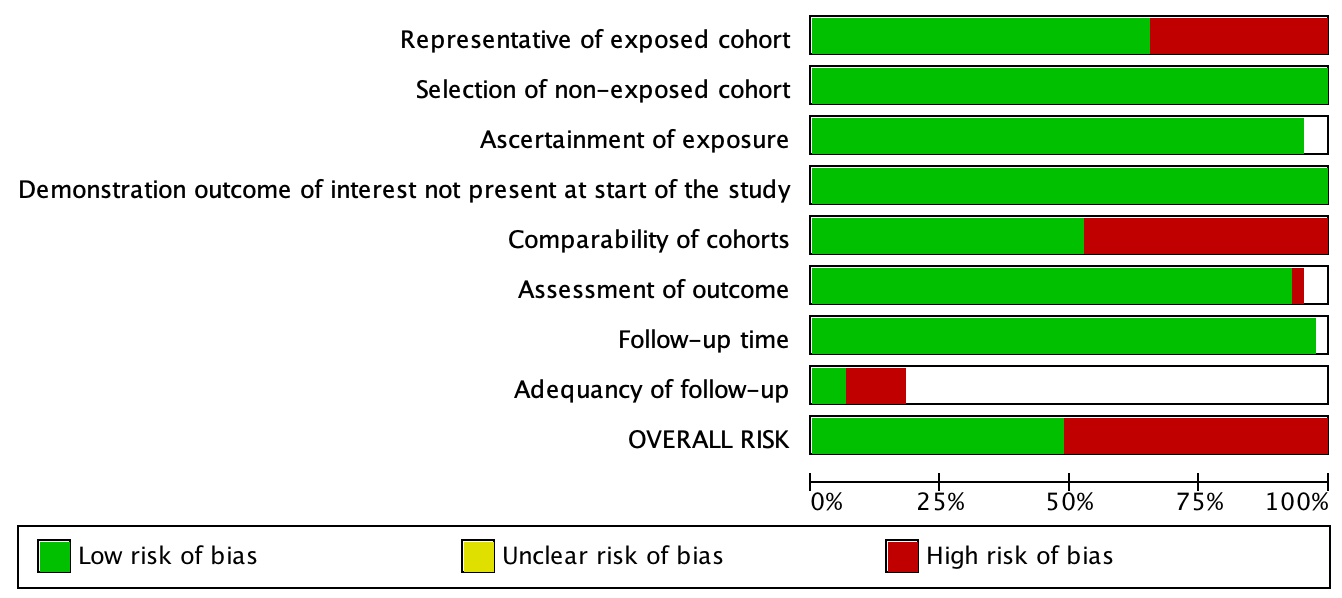

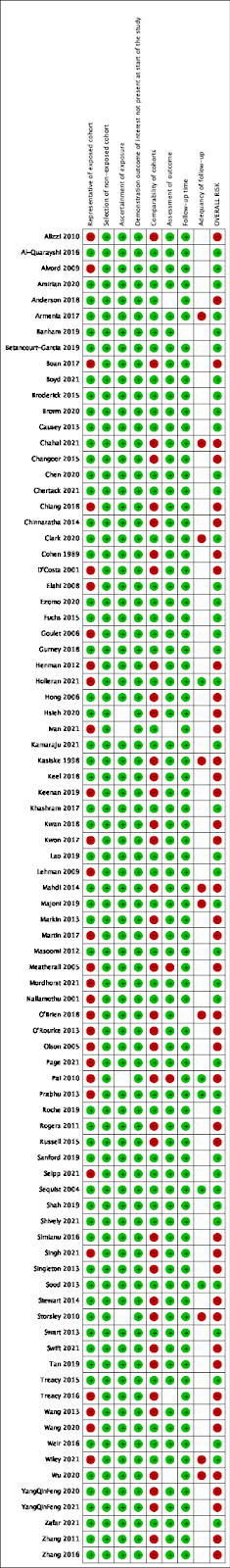
