## Supplementary material for "Post-operative morbidity and mortality in Indigenous Peoples: A scoping review and meta-analysis": S2 Appendix

**S2 Appendix: Search terms**

**OVID Medline ® ALL <1946 to December 23, 2021>**

Indians,north american.mp. [mp=title, abstract, original title, name of substance word, subject heading word, floating sub-heading word, keyword heading word, organism supplementary concept word, protocol supplementary concept word, rare disease supplementary concept word, unique identifier, synonyms] OR exp Inuits/ OR exp Health Services, Indigenous/ OR exp Indigenous Peoples/ OR (Athapaskan or Saulteaux).mp. OR (Wakashan or Cree or Dene or Inuit or Inuk or Inuvialuit* or Haida).mp. OR maori.mp. [mp=title, abstract, original title, name of substance word, subject heading word, floating sub-heading word, keyword heading word, organism supplementary concept word, protocol supplementary concept word, rare disease supplementary concept word, unique identifier, synonyms] OR maori.mp. or exp Oceanic Ancestry Group/ OR exp Alaskan Natives/ OR eskimo.mp. OR (Ktunaxa or Assiniboine or Akwesasne or Iroquois).mp. OR (Inupiat* or Miawpukek or Mushkegowuk or Naskapi or Oji-Cree or Opaskwayak).mp. OR polynesian*.mp. OR native hawaiian.mp. OR (Pauktuutit or Qikiqtani or Wikwemikong or Tsimshian or Gitsxan or Nisga'a or Haisla or Heiltsuk or Oweenkeno or Kwakwaka'wakw or Nuu chah nulth or Tsilhqot'in).mp. OR (Dakelh or Wet'suwet'en or Sekani or Dunne-za or Dene or Tahltan or Kaska or Tagish or Tutchone or Nuxalk or Salish or Stl'atlimc or Nlaka'pamux or Okanagan or Sec wepmc or Tlingit or Anishinaabe or Blackfoot or Nakoda or Tasttine or Tsuu T'inia or Gwich'in or Han or Tagish or Tutchone or Algonquin or Nipissing or Ojibwa or Potawatomi or Innu or Maliseet or Mi'kmaq or Micmac).mp. OR (Passamaquoddy or Haudenosaunee or Cayuga or Mohawk or Oneida or Onodaga or Seneca or Tuscarora or Wyandot or Aboriginal* or Indigenous* or Metis or red road or "on reserve" or off-reserve or First Nation or First Nations or Amerindian or (urban adj3 (Indian* or Native* or Aboriginal*))).mp. OR (autochtone* or (Native* adj1 (man or men or women or woman or boy* or girl* or adolescent* or youth or youths or person* or adult or people* or Indian* or Nation or tribe* or tribal or band or bands))).mp. OR Indians, South American/ OR quechua*.mp. OR chiquitano*.mp. OR mestizo.mp. OR guaranies.mp. OR amerid*.mp.

AND

exp surgery/ or surgical patient/ or (surger* or surgical or operation* or operative or transplant*).mp. or exp hip fracture/ or perioperative period/ or preoperative care/ or preoperative period/ or intraoperative period/ or postoperative period/ or (perioperative* or peri-operative*).mp. or (preoperative* or pre-operative*).mp. or (intraoperative* or intra-operative*).mp. or (postoperative* or post-operative* or postsurg* or post-surg*).mp. or postoperative complication/ or (obstetric* or cesarean or c-section or section or childbirth or child birth).mp. or transplant.mp. OR outcome*.mp.

AND outcome*.mp.

**EMBASE <1974 to 2021 December 23>**

exp American Indians/ or exp Tribes/ or exp Indigenous Populations/ or exp Alaska Natives/ or (Athapaskan or Saulteaux).mp. or exp Inuit/ or (Wakashan or Cree or Dene or Inuit or Inuk or Inuvialuit* or Haida).mp. or maori.mp. or exp Pacific Islanders/ or eskimo.mp. or (Ktunaxa or Assiniboine or Akwesasne or Iroquois).mp. or (Inupiat* or Miawpukek or Mushkegowuk or Naskapi or Oji-Cree or Opaskwayak).mp. or polynesian.mp. or exp Hawaii Natives/ or (Pauktuutit or Qikiqtani or Wikwemikong or Tsimshian or Gitsxan or Nisga'a or Haisla or Heiltsuk or Oweenkeno or Kwakwaka'wakw or Nuu chah nulth or Tsilhqot'in).mp. OR exp American Indian/ OR (Dakelh or Wet'suwet'en or Sekani or Dunne-za or Dene or Tahltan or Kaska or Tagish or Tutchone or Nuxalk or Salish or Stl'atlimc or Nlaka'pamux or Okanagan or Sec wepmc or Tlingit or Anishinaabe or Blackfoot or Nakoda or Tasttine or Tsuu T'inia or Gwich'in or Han or Tagish or Tutchone or Algonquin or Nipissing or Ojibwa or Potawatomi or Innu or Maliseet or Mi'kmaq or Micmac or (Passamaquoddy or Haudenosaunee or Cayuga or Mohawk or Oneida or Onodaga or Seneca or Tuscarora or Wyandot or Aboriginal* or Indigenous* or Metis or red road or "on reserve" or off-reserve or First Nation or First Nations or Amerindian or (urban adj3 (Indian* or Native* or

Aboriginal*))) or (autochtone* or (Native* adj1 (man or men or women or woman or boy* or girl* or adolescent* or youth or youths or person* or adult or people* or Indian* or Nation or tribe* or tribal or band or bands)))).mp. OR exp "Quechua (people)"/ OR chiquitano.mp. OR exp Mestizo/ OR exp "Guarani (people)"/ OR inca.mp. OR amerid*.mp. OR exp Metis/ OR exp Native Hawaiian/

AND

exp surgery/ or surgical patient/ or (surger* or surgical or operation* or operative or transplant*).mp. or exp hip fracture/ or perioperative period/ or preoperative care/ or preoperative period/ or intraoperative period/ or postoperative period/ or (perioperative* or peri-operative*).mp. or (preoperative* or pre-operative*).mp. or (intraoperative* or intra-operative*).mp. or (postoperative* or post-operative* or postsurg* or post-surg*).mp. or postoperative complication/ or (obstetric* or cesarean or c-section or section or childbirth or child birth).mp. or transplant.mp.

AND

exp adverse outcome/ or exp critical care outcome/ or exp treatment outcome/ or exp clinical outcome/ OR outcome*.mp.

**CINAHL**

( ( ( (MH "Eskimos") OR (MH "Native Americans") OR (MH "Indigenous Peoples+") or (MH "Health Services, Indigenous") or (MH "Indigenous Health") or (MH “Ethnopharmacology”) or Athapaskan or Saulteaux or Wakashan or Cree or Dene or Inuit or Alaskan native or Inuk or Inuvialuit* or Haida or Ktunaxa or Assiniboine or Akwesasne or 3,210 Iroquois.mp. Inupiat* or Miawpukek or Mushkegowuk. or Naskapi or Oji-Cree or Opaskwayak or Pauktuutit or Qikiqtani or Wikwemikong or Tsimshian or Gitsxan or “Nisga'a” or Haisla or Heiltsuk or Oweenkeno or “Kwakwaka'wakw” or “Nuu chah nulth” or “Tsilhqot'in” or Dakelh or Wet'suwet'en” or Sekani or “Dunne-za” or Dene or Tahltan or Kaska or Tagish or Tutchone or Nuxalk or Salish or “Stl'atlimc” or “Nlaka'pamux” or Okanagan or “Sec wepmc” or Tlingit or Anishinaabe or Blackfoot or Nakoda or Tasttine or “Tsuu T'inia” or “Gwich'in” or Han or Tagish or Tutchone or Algonquin or Nipissing or Ojibwa or Potawatomi or Innu or Maliseet or “Mi'kmaq” or Micmac or “Mic mac” Passamaquoddy or Haudenosaunee or Cayuga or Mohawk or Oneida or Onodaga or Seneca or Tuscarora or Wyandot or Aboriginal* or Indigenous* or Metis or “red road” or "on reserve" or “off reserve” or “First Nation” or “First Nations” or Amerindian or (urban N3 (Indian* or Native* or Aboriginal*) or Maori) ) OR ( (MH "Indians, South American") OR (MH "quechua") OR (MH "Pacific Islanders") or (MH "amerid*") or (MH "chiquitano") or (MH "mesitzo") or (MH "guaranies") )

AND

(obstetric* or c?esarean or c-section) OR AB ((obstetric* or c?esarean or c-section) OR ( (postoperative* or post-operative* or postsurg* or post-surg*) ) OR AB ((postoperative* or post-operative* or postsurg* or post-surg*)) OR ( (intraoperative* or intra-operative*). ) OR AB ( (intraoperative* or intraoperative*). ) OR ( (preoperative* or pre-operative*) ) OR AB ( (preoperative* or preoperative*) ) OR ( (perioperative* or peri-operative*) ) OR AB ( (perioperative* or perioperative*)) OR (MH "Postoperative Period") OR (MH "Postoperative Complications+") OR (MH "Intraoperative Period") OR (MH "Preoperative Period+") OR ( (surger* or surgical or operation or operative or transplant* ) OR AB ((surger* or surgical or operation or operative or transplant*) OR (MH "Surgery, Operative+") OR (MH "Surgical Patients")

AND

MH "outcome*"

**APA PsycInfo <1806 to December Week 3 2021>**

exp American Indians/ or exp Tribes/ or exp Indigenous Populations/ or exp Alaska Natives/ OR exp Inuit/ OR (Athapaskan or Saulteaux).mp. OR (Wakashan or Cree or Dene or Inuit or Inuk or Inuvialuit* or Haida).mp. OR maori.mp. OR exp Pacific Islanders/ OR eskimo.mp. OR (Ktunaxa or Assiniboine or Akwesasne or Iroquois).mp. OR (Inupiat* or Miawpukek or Mushkegowuk or Naskapi or Oji-Cree or Opaskwayak).mp. OR polynesian.mp. OR exp Hawaii Natives/ OR (Pauktuutit or Qikiqtani or Wikwemikong or Tsimshian or Gitsxan or Nisga'a or Haisla or Heiltsuk or Oweenkeno or Kwakwaka'wakw or Nuu chah nulth or Tsilhqot'in).mp. OR (Dakelh or Wet'suwet'en or Sekani or Dunne-za or Dene or Tahltan or Kaska or Tagish or Tutchone or Nuxalk or Salish or Stl'atlimc or Nlaka'pamux or Okanagan or Sec wepmc or Tlingit or Anishinaabe or Blackfoot or Nakoda or Tasttine or Tsuu T'inia or Gwich'in or Han or Tagish or Tutchone or Algonquin or Nipissing or Ojibwa or Potawatomi or Innu or Maliseet or Mi'kmaq or Micmac).mp. OR (Passamaquoddy or Haudenosaunee or Cayuga or Mohawk or Oneida or Onodaga or Seneca or Tuscarora or Wyandot or Aboriginal* or Indigenous* or Metis or red road or "on reserve" or off-reserve or First Nation or First Nations or Amerindian or (urban adj3 (Indian* or Native* or Aboriginal*))).mp. OR (autochtone* or (Native* adj1 (man or men or women or woman or boy* or girl* or adolescent* or youth or youths or person* or adult or people* or Indian* or Nation or tribe* or tribal or band or bands))).mp. OR quechua.mp. OR south american natives.mp. OR chiquitano*.mp. OR mestizo.mp. OR guaranies.mp. OR amerid*.mp.

AND

exp surgery/ or surgical patient/ or (surger* or surgical or operation* or operative or transplant*).mp. or perioperative period/ or preoperative care/ or preoperative period/ or intraoperative period/ or postoperative period/ or (perioperative* or peri-operative*).mp. or (preoperative* or pre-operative*).mp. or (intraoperative* or intra-operative*).mp. or (postoperative* or post-operative* or postsurg* or post-surg*).mp. or postoperative complication/ or (obstetric* or cesarean or c-section or section or childbirth or child birth).mp. or transplant.mp.

**SocIndex**

"( (obstetric* or c?esarean or c-section) OR AB ((obstetric* or c?esarean or c-section) OR ( (postoperative* or post-operative* or postsurg* or post-surg*) ) OR AB ((postoperative* or post-operative* or postsurg* or post-surg*) ) OR ( (intraoperative* or intra-operative*). ) OR AB ( (intraoperative* or intraoperative*). ) OR ( (preoperative* or pre-operative*) ) OR AB ( (preoperative* or preoperative*) ) OR ( (perioperative* or peri-operative*) ) OR AB ( (perioperative* or perioperative*)) OR (MH "Postoperative Period") OR (MH "Postoperative Complications+") OR (MH "Intraoperative Period") OR (MH "Preoperative Period+") OR ( (surger* or surgical or operation or operative or transplant* ) OR AB ((surger* or surgical or operation or operative or transplant*) OR (MH "Surgery, Operative+") OR (MH "Surgical Patients") )

AND

( ( ( ( (MH "Eskimos") OR (MH "Native Americans") OR (MH "Indigenous Peoples+") or (MH "Health Services, Indigenous") or (MH "Indigenous Health") or (MH “Ethnopharmacology”) or Athapaskan or Saulteaux or Wakashan or Cree or Dene or Inuit or Alaskan native or Inuk or Inuvialuit* or Haida or Ktunaxa or Assiniboine or Akwesasne or 3,210 Iroquois.mp. Inupiat* or Miawpukek or Mushkegowuk. or Naskapi or Oji-Cree or Opaskwayak or Pauktuutit or Qikiqtani or Wikwemikong or Tsimshian or Gitsxan or “Nisga'a” or Haisla or Heiltsuk or Oweenkeno or “Kwakwaka'wakw” or “Nuu chah nulth” or “Tsilhqot'in” or Dakelh or Wet'suwet'en” or Sekani or “Dunne-za” or Dene or Tahltan or Kaska or Tagish or Tutchone or Nuxalk or Salish or “Stl'atlimc” or “Nlaka'pamux” or Okanagan or “Sec wepmc” or Tlingit or Anishinaabe or Blackfoot or Nakoda or Tasttine or “Tsuu T'inia” or “Gwich'in” or Han or Tagish or Tutchone or Algonquin or Nipissing or Ojibwa or Potawatomi or Innu or Maliseet or “Mi'kmaq” or Micmac or “Mic mac” Passamaquoddy or Haudenosaunee or Cayuga or Mohawk or Oneida or Onodaga or Seneca or Tuscarora or Wyandot or Aboriginal* or Indigenous* or Metis or “red road” or "on reserve" or “off reserve” or “First Nation” or “First Nations” or Amerindian or (urban N3 (Indian* or Native* or Aboriginal*) or Maori) ) OR ( (MH "Indians, South American") OR (MH "quechua") OR (MH "Pacific Islanders") or (MH

"amerid*") or (MH "chiquitano") or (MH "mesitzo") or (MH "guaranies") ) ) Apply equivalent subjects on 2021-12-26 06:20 AM"

**Global Health <1910 to 2021 Week 50>**

exp indigenous people/ OR American indians/ OR Alaska Natives/ OR Inuit/ OR (Athapaskan or Saulteaux).mp. OR (Wakashan or Cree or Dene or Inuit or Inuk or Inuvialuit* or Haida).mp. OR maori.mp. OR Pacific Islanders/ OR eskimo.mp. OR (Ktunaxa or Assiniboine or Akwesasne or Iroquois).mp. OR (Inupiat* or Miawpukek or Mushkegowuk or Naskapi or Oji-Cree or Opaskwayak).mp. OR polynesian.mp. OR Hawaii Natives.mp. OR (Pauktuutit or Qikiqtani or Wikwemikong or Tsimshian or Gitsxan or Nisga'a or Haisla or Heiltsuk or Oweenkeno or Kwakwaka'wakw or Nuu chah nulth or Tsilhqot'in).mp. OR (Dakelh or Wet'suwet'en or Sekani or Dunne-za or Dene or Tahltan or Kaska or Tagish or Tutchone or Nuxalk or Salish or Stl'atlimc or Nlaka'pamux or Okanagan or Sec wepmc or Tlingit or Anishinaabe or Blackfoot or Nakoda or Tasttine or Tsuu T'inia or Gwich'in or Han or Tagish or Tutchone or Algonquin or Nipissing or Ojibwa or Potawatomi or Innu or Maliseet or Mi'kmaq or Micmac).mp. OR (Passamaquoddy or Haudenosaunee or Cayuga or Mohawk or Oneida or Onodaga or Seneca or Tuscarora or Wyandot or Aboriginal* or Indigenous* or Metis or red road or "on reserve" or off-reserve or First Nation or First Nations or Amerindian or (urban adj3 (Indian* or Native* or Aboriginal*))).mp. OR (autochtone* or (Native* adj1 (man or men or women or woman or boy* or girl* or adolescent* or youth or youths or person* or adult or people* or Indian* or Nation or tribe* or tribal or band or bands))).mp. OR aboriginal.mp. or aborigines.sh. OR quechua.mp. OR south american natives.mp. OR chiquitano*.mp. OR mestizo.mp. OR guaranies.mp. OR amerid*.mp.

AND

exp surgery/ or surgical patient/ or (surger* or surgical or operation* or operative or transplant*).mp. or perioperative period/ or preoperative care/ or preoperative period/ or intraoperative period/ or postoperative period/ or (perioperative* or peri-operative*).mp. or (preoperative* or pre-operative*).mp. or (intraoperative* or intra-operative*).mp. or (postoperative* or post-operative* or postsurg* or post-surg*).mp. or postoperative complication/ or (obstetric* or cesarean or c-section or section or childbirth or child birth).mp. or transplant.mp.

**Proquest Dissertations and Theses Global**

((ti((aboriginal OR "first nation" OR "first nations" OR inuit OR metis OR "indigenous canadians" OR "native people" OR "native peoples" OR "on-reserve" OR "off-reserve")) OR ab((aboriginal OR "firstnation" OR "first nations" OR inuit OR Alaskan native OR Maori OR metis OR "indigenous canadians" OR "native people" OR "native peoples" OR "on-reserve" OR "off-reserve"))) OR noft((Algonquin OR Aleut* OR Anishinabek OR Anishnabek OR Chipewyan OR Cree OR Dene OR eskimo* OR Gitskan OR Huron OR Innu OR Inuktitut OR Maori OR Amerid* OR "Indians, South America*" OR Quechua OR "Pacific Islander*" OR Chiquitano OR Mesitzo OR Guaranie* OR Inuk OR Inupiat* OR Iqaluit OR Iroquois OR Kalaallit* OR "Kawawachikamach Québec" OR Kahnawa:ke OR Kitikmeot OR Kitimat OR Kivalliq OR Kwakiutl OR Manitoulin OR Metis OR Miawpukek OR Micmac OR Mi’kmaq OR Mohawk OR Mushkegowuk OR Naskapi OR Nisga'a OR Nakoda OR Nakota OR Oji-Cree OR Ojibway OR Oki OR Opaskwayak OR Pauktuutit OR Qikiqtani OR Qayuqtuvik OR "Rankin Inlet" OR Sekon OR Sioux OR Tungasugit OR Tuttarvingat OR "Vuntut Gwitchin")))

AND

(ti(surger* OR surgical OR operation OR operative OR transplant*) OR ti(obstetric* OR cesarean OR caesarean OR c-section) OR (ab(perioperative* OR peri-operative) OR ab(preoperative* OR pre-operative*) OR ab(intraoperative* OR intra-operative*) OR ab(postoperative* OR post-operative* OR postsurg* OR post-surg*)) OR su((surger* OR surgical OR operation OR operative OR transplant*)))
