## Supplementary material for "Post-operative morbidity and mortality in Indigenous Peoples: A scoping review and meta-analysis": S3 Appendix

**S3 Appendix: References of Included Studies**

a. **Included in Meta-Analysis**

Al-Qurayshi Z, Randolph GW, Srivastav S, Aslam R, Friedlander P, Kandil E. Outcomes in thyroid surgery are affected by racial, economic, and healthcare system demographics. *The Laryngoscope*. 2016;126(9):2194-2199. doi:10.1002/lary.25871

Alizzi AM, Knight JL, Tully PJ. Surgical challenges in rheumatic heart disease in the Australian indigenous population. *Heart, Lung and Circulation*. 2010;19(5-6):295-298. doi:10.1016/j.hlc.2010.02.010

Amirian H, Torquati A, Omotosho P. Racial disparity in 30-day outcomes of metabolic and Bariatric Surgery. *Obesity Surgery*. 2019;30(3):1011-1020. doi:10.1007/s11695-019-04282-9

Armenia SJ, Pentakota SR, Merchant AM. Socioeconomic factors and mortality in emergency general surgery: Trends over a 20-year period. *Journal of Surgical Research*. 2017;212:178-186. doi:10.1016/j.jss.2017.01.015

Alvord LA, Henderson WG, Benton K, Buchwald D. Surgical Outcomes in American Indian veterans: A closer look. *Journal of the American College of Surgeons*. 2009;208(6). doi:10.1016/j.jamcollsurg.2009.02.058

Banham D, Roder D, Eckert M, Howard NJ, Canuto K, Brown A. Cancer treatment and the risk of cancer death among Aboriginal and non-Aboriginal South Australians: Analysis of a matched Cohort Study. *BMC Health Services Research*. 2019;19(1). doi:10.1186/s12913-019-4534-y

Betancourt-Garcia MM, Vatcheva K, Gupta PK, et al. The effect of Hispanic ethnicity on surgical outcomes: An analysis of the NSQIP database. *The American Journal of Surgery*. 2019;217(4):618-633. doi:10.1016/j.amjsurg.2018.10.004

Boan P, Swaminathan R, Irish A. Infectious complications in indigenous renal transplant recipients in Western Australia. *Internal Medicine Journal*. 2017;47(6):648-655. doi:10.1111/imj.13450

Brown O, Geynisman-Tan J, Gillingham A, et al. Minimizing risks in minimally invasive surgery: Rates of surgical site infection across subtypes of laparoscopic hysterectomy. *Journal of Minimally Invasive Gynecology*. 2020;27(6). doi:10.1016/j.jmig.2019.10.015

Causey MW, McVay D, Hatch Q, et al. The impact of race on outcomes following emergency surgery: An American College of Surgeons National Surgical Quality Improvement Program Assessment. *The American Journal of Surgery*. 2013;206(2):172-179. doi:10.1016/j.amjsurg.2012.11.022

Chahal D, Marquez V, Hussaini T, et al. End stage liver disease etiology & transplantation referral outcomes of major ethnic groups in British Columbia, Canada. *Medicine*. 2021;100(42). doi:10.1097/md.0000000000027436

Changoor NR, Ortega G, Ekladios M, Zogg CK, Cornwell EE, Haider AH. Racial disparities in surgical outcomes: Does the level of resident surgeon play a role? *Surgery*. 2015;158(2):547-555. doi:10.1016/j.surg.2015.03.046

Chen Y, Chang D. Variation in the risk of venous thromboembolism after major abdominal surgeries across racial groups. *Gastroenterology*. 2020;158(6). doi:10.1016/s0016-5085(20)31915-6

Chertack N, Baky F, Samplaski MK, Vij SC, Bakare T. The impact of race and gender on 30-day urologic surgery complications. *Urology*. 2021;162:77-83. doi:10.1016/j.urology.2021.05.023

Chiang N, Jain JK, Hulme KR, Vasudevan T. Epidemiology and outcomes of abdominal aortic aneurysms in New Zealand: A 15-year experience at a Regional Hospital. *Annals of Vascular Surgery*. 2018;46:274-284. doi:10.1016/j.avsg.2017.07.006

Cohen MM, Young TK, Hammarstrand KM. Ethnic variation in cholecystectomy rates and outcomes, Manitoba, Canada, 1972-84. *American Journal of Public Health*. 1989;79(6):751-755. doi:10.2105/ajph.79.6.751

D'Costa R, Ruygrok PN, Coverdale HA, et al. Cardiac transplantation in New Zealand Maori and Polynesian patients. *Heart, Lung and Circulation*. 2000;9(3). doi:10.1046/j.1443-9506.2000.08849.x

Elahi M, Matata B, Yii M. Ethnicity and adverse operative outcomes among Australian patients undergoing first-time isolated coronary artery bypass graft surgery. *International Surgery*. 2008;93(6):358-65. PMID: 20085046

Goulet S, Trepman E, MMath MC, et al. Revascularization for peripheral vascular disease in Aboriginal and non-Aboriginal patients. *Journal of Vascular Surgery*. 2006;43(4):735-741. doi:10.1016/j.jvs.2005.11.058

Gurney JK, Stanley J, Rumball-Smith J, York S, Sarfati D. Postoperative death after lower-limb amputation in a national prevalent cohort of patients with diabetes. *Diabetes Care*. 2018;41(6):1204-1211. doi:10.2337/dc17-2557

Henman K, Gordon CL, Gardiner T, et al. Surgical site infections following caesarean section at Royal Darwin Hospital, Northern Territory. *Healthcare infection*. 2012;17(2):47-51. doi:10.1071/hi11027

Holleran TJ, Napolitano MA, LaPiano JB, et al. Racial disparities in 30-day outcomes after colorectal surgery in an integrated healthcare system. *Journal of Gastrointestinal Surgery*. 2021;26(2):433-443. doi:10.1007/s11605-021-05151-6

Hsieh Y-C, Shah HR, Balasubramaniam P. The association of race with outcomes among parturients undergoing cesarean section with perioperative epidural catheter placement: A nationwide analysis. *Cureus*. 2020. doi:10.7759/cureus.6652

Ivan E, Martinsen B, Igyarto Z, Sublett T, Nachimuthu S. Peripheral artery disease in vulnerable patient populations: Outcomes of orbital atherectomy in Native Americans compared to Non-Native Americans. A single-center experience in rural Oklahoma. *Cardiovascular Revascularization Medicine*. 2021;22:71-77. doi:10.1016/j.carrev.2020.06.008

Kamaraju A, Feinn R, Myrick K, Halawi MJ. Total versus Unicondylar Knee Arthroplasty: Does race play a role in the treatment selection? *Journal of Racial and Ethnic Health Disparities*. 2021. doi:10.1007/s40615-021-01120-6

Kasiske BL, Chakkera H. Successful renal transplantation in American Indians. *Transplantation*. 1998;66(2):209-214. doi:10.1097/00007890-199807270-00012

Keenan NM, Newland RF, Baker RA, Rice GD, Bennetts JS. Outcomes of Redo Valve Surgery in indigenous Australians. *Heart, Lung and Circulation*. 2019;28(7):1102-1111. doi:10.1016/j.hlc.2018.05.198

Khashram M, Pitama S, Williman JA, Jones GT, Roake JA. Survival disparity following abdominal aortic aneurysm repair highlights inequality in ethnic and socio-economic status. *European Journal of Vascular and Endovascular Surgery*. 2017;54(6):689-696. doi:10.1016/j.ejvs.2017.08.018

Kwan JM, Hajjiri Z, Chen YF, Metwally A, Perkins DL, Finn PW. Donor and recipient ethnicity impacts renal graft adverse outcomes. *Journal of Racial and Ethnic Health Disparities*. 2017;5(5):1003-1013. doi:10.1007/s40615-017-0447-9

Kwon HJ, Morton RP. Ethnic disparities in thyroid surgery outcomes in New Zealand. *ANZ Journal of Surgery*. 2015;87(7-8):610-614. doi:10.1111/ans.13142

Lao C, Lees D, Patel S, White D, Lawrenson R. Geographical and ethnic differences of osteoarthritis-associated hip and knee replacement surgeries in New Zealand: A population-based cross-sectional study. *BMJ Open*. 2019;9(9). doi:10.1136/bmjopen-2019-032993

Lehman SJ, Baker RA, Aylward PE, Knight JL, Chew DP. Outcomes of cardiac surgery in indigenous Australians. *Medical Journal of Australia*. 2009;190(10):588-593. doi:10.5694/j.1326-5377.2009.tb02573.x

Mahdi H, Schlick CJ, Kowk L-L, Moslemi-Kebria M, Michener C. Endometrial cancer in Asian and American Indian/Alaskan Native women: Tumor characteristics, treatment and outcome compared to non-Hispanic white women. *Gynecologic Oncology*. 2014;132(2):443-449. doi:10.1016/j.ygyno.2013.11.028

Majoni SW, Ullah S, Collett J, Hughes JT, McDonald S. Weight change trajectories in Aboriginal and Torres Strait Islander Australians after Kidney Transplantation: A cohort analysis using the Australia and New Zealand dialysis and Transplant Registry (ANZDATA). *BMC Nephrology*. 2019;20(1). doi:10.1186/s12882-019-1411-1

Markin A, Habermann EB, Zhu Y, et al. Cancer surgery among American Indians. *JAMA Surgery*. 2013;148(3):277. doi:10.1001/jamasurg.2013.1423

Martin JR, Wang TY, Loriaux D, et al. Race as a predictor of postoperative hospital readmission after Spine Surgery. *Journal of Clinical Neuroscience*. 2017;46:21-25.doi:10.1016/j.jocn.2017.08.015

Meatherall BL, Garrett MR, Kaufert J, et al. Disability and quality of life in Canadian aboriginal and Non-Aboriginal diabetic lower-extremity amputees. *Archives of Physical Medicine and Rehabilitation*. 2005;86(8):1594-1602. doi:10.1016/j.apmr.2004.11.026

Nallamothu BK, Saint S, Saha S, Fendrick AM, Kelley K, Ramsey SD. Coronary artery bypass grafting in Native Americans. *Journal of General Internal Medicine*. 2001;16(8):554-559. doi:10.1046/j.1525-1497.2001.016008554.x

O'Brien J, Saxena A, Reid CM, et al. Thirty-day outcomes in indigenous Australians following coronary artery bypass grafting. *Internal Medicine Journal*. 2018;48(7):780-785. doi:10.1111/imj.13790

O'Rourke S, Steffen C, Raulli A, Tulip F. Diabetic major amputation in Far North Queensland 1998-2008: What is the gap for indigenous patients? *Australian Journal of Rural Health*. 2013;21(5):268-273. doi:10.1111/ajr.12044

Page S, Yong MS, Saxena P, Yadav S. Outcomes in dialysis-dependent indigenous and Non-Indigenous patients undergoing cardiac surgery at Townsville University Hospital. *Heart, Lung and Circulation*. 2021;30(8):1200-1206. doi:10.1016/j.hlc.2021.02.013

Pai V, Pai V, Wright S. Differences in outcome between Maori and Caucasian patients undergoing total joint arthroplasty for osteoarthritis. *Journal of Orthopaedic Surgery*. 2010;18(2):195-197. doi:10.1177/230949901001800212

Prabhu A, Tully PJ, Bennetts JS, Tuble SC, Baker RA. The morbidity and mortality outcomes of Indigenous Australian peoples after isolated coronary artery bypass graft surgery: The influence of geographic remoteness. *Heart, Lung and Circulation*. 2013;22(8):599-605. doi:10.1016/j.hlc.2013.01.003

Roche M, Law Tyee, Sultan AA, et al. Racial disparities in revision total knee arthroplasty: Analysis of 125,901 patients in National US Private Payer Database. *Journal of Racial and Ethnic Health Disparities*. 2018;6(1):101-109. doi:10.1007/s40615-018-0504-z

Rogers NM, Lawton PD, Jose MD. Plasma cell infiltrates and renal allograft outcomes in indigenous and non-indigenous people of the Northern Territory of Australia. *Nephrology*. 2011;16(8):777-783. doi:10.1111/j.1440-1797.2011.01487.x

Russell EA, Tran L, Baker RA, et al. A review of outcome following valve surgery for rheumatic heart disease in Australia. *BMC Cardiovascular Disorders*. 2015;15(1). doi:10.1186/s12872-015-0094-1

Sanford Z, Taylor H, Fiorentino A, et al. Racial disparities in surgical outcomes after spine surgery: An ACS-NSQIP analysis. *Global Spine Journal*. 2018;9(6):583-590. doi:10.1177/2192568218811633

Seipp R, Zhang N, Nair SS, et al. Patient and allograft outcomes after kidney transplant for the indigenous patients in the United States. *PLOS ONE*. 2021;16(2). doi:10.1371/journal.pone.0244492

Simianu VV, Morris AM, Varghese TK, et al. Evaluating disparities in inpatient surgical cancer care among American Indian/alaska native patients. *The American Journal of Surgery*. 2016;212(2):297-304. doi:10.1016/j.amjsurg.2015.10.030

Singh TP, Moxon JV, Meehan MT, Jones R, Cadet-James Y, Golledge J. Major amputation rates and outcomes for Aboriginal and Torres Strait Islander and non-Indigenous people in North Queensland Australia between 2000 and 2015. *BMC Endocrine Disorders*. 2021;21(1). doi:10.1186/s12902-021-00764-z

Sood MM, Tangri N, Komenda P, et al. Incidence, secular trends and outcomes of cardiac surgery in Aboriginal Peoples. *Canadian Journal of Cardiology*. 2013;29(10). doi:10.1016/j.cjca.2013.07.357

Stewart FR, Ruygrok PN, Gibbs HC, et al. Outcome following heart transplantation in New Zealand Maori. *Heart, Lung and Circulation*. 2014;23(4):353-356. doi:10.1016/j.hlc.2013.11.014

Storsley LJ, Young A, Rush DN, et al. Long-term medical outcomes among Aboriginal Living Kidney Donors. *Transplantation*. 2010;90(4):401-406. doi:10.1097/tp.0b013e3181e6e79b

Swart EM, Sarfati D, Cunningham R, Dennett E, Signal V, Gurney J, Stanley J. Ethnicity and rectal cancer management in New Zealand. *New Zealand Medical Journal*. 2013;126(1384):42-52. PMID: 24162629

Swift K, Thompson F, Roeder L, Choy KT, McDonald M, Costa A. Appendicitis in Far North Queensland: A new take on an old story. *ANZ Journal of Surgery*. 2021;92(1-2):114-120. doi:10.1111/ans.17404

Tan T-W, Shih C-D, Concha-Moore KC, et al. Disparities in outcomes of patients admitted with diabetic foot infections. *PLOS ONE*. 2019;14(2). doi:10.1371/journal.pone.0211481

Treacy PJ, Chatfield MD, Bessell J. Is gastric banding appropriate in indigenous or remote-dwelling persons? *Obesity Surgery*. 2016;26(8):1728-1734. doi:10.1007/s11695-015-1993-z

Treacy PJ, North JB, Rey-Conde T, Allen J, Ware RS. Outcomes from the Northern Territory Audit of surgical mortality: Aboriginal deaths. *ANZ Journal of Surgery*. 2015;85(1-2):11-15. doi:10.1111/ans.12896

Wang TKM, Ramanathan T, Stewart RAH, Crengle S, Gamble GD, White HD. Maori have worse outcomes after coronary artery bypass grafting than Europeans: Another example of ethnic disparity in cardiovascular disease. *Heart, Lung and Circulation*. 2013;22(7):588. doi:10.1016/j.hlc.2013.04.095

Wang TKM, Wei D, Evans T, Ramanathan T, Haydock D. Comparison of characteristics and outcomes for type A aortic dissection surgery by Māori, Pasifika or other ethnicities. *New Zealand Medical Journal*. 2020;133(1514):33-40. PMID: 32379737

Weir K, Supramaniam R, Gibberd A, Dillon A, Armstrong BK, O'Connell DL. Comparing colorectal cancer treatment and survival for Aboriginal and non‐aboriginal people in New South Wales. *Medical Journal of Australia*. 2016;204(4):156-156. doi:10.5694/mja15.01153

Wiley HRL, Varilek BM, Saucedo-Crespo H, et al. Kidney Transplant Outcomes in indigenous people of the Northern Great Plains of the United States. *Transplantation Proceedings*. 2021;53(6):1872-1879. doi:10.1016/j.transproceed.2021.05.003

Wu J, Ebrahim AK. Ethnic disparities for thyroid surgery. *ANZ Journal of Surgery*. 2020;90(12):2527-2531. doi:10.1111/ans.16410

Yang Q, Lin Z, Yang S, Wang P, Chen R, Wang J. Incidence and risk factors of in‐hospital prosthesis‐related complications following total knee arthroplasty: A retrospective nationwide inpatient sample database study. *Orthopaedic Surgery*. 2021;13(5):1579-1586. doi:10.1111/os.13008

Yang Q, Wang J, Xu Y, Chen Y, Lian Q, Zhang Y. Incidence and risk factors of in-hospital prosthesis-related complications following total hip arthroplasty: A retrospective nationwide inpatient sample database study. *International Orthopaedics*. 2020;44(11):2243-2252. doi:10.1007/s00264-020-04682-y

Zhang M, Uhanova J, Minuk GY. Liver transplant outcomes in a canadian first nations population. *Canadian Journal of Gastroenterology*. 2011;25(6):307-310. doi:10.1155/2011/986945

Zhang W, Lyman S, Boutin-Foster C, et al. Racial and ethnic disparities in utilization rate, hospital volume, and perioperative outcomes after total Knee Arthroplasty. *Journal of Bone and Joint Surgery*. 2016;98(15):1243-1252. doi:10.2106/jbjs.15.01009

b. **Included Only in Narrative Synthesis**

Anderson E, Glogoza M, Bettenhausen A, et al. Disparities in cardiovascular risk factors in northern plains American Indians undergoing coronary artery bypass grafting. *Health Equity*. 2018;2(1):152-160. doi:10.1089/heq.2018.0021

Boyd BAJ, Winkelman WD, Mishra K, Vittinghoff E, Jacoby VL. Racial and ethnic differences in reconstructive surgery for apical vaginal prolapse. *American Journal of Obstetrics and Gynecology*. 2021;225(4). doi:10.1016/j.ajog.2021.05.002

Broderick RC, Fuchs HF, Harnsberger CR, et al. Increasing the value of healthcare: Improving mortality while reducing cost in bariatric surgery. *Obesity Surgery*. 2015;25(12):2231-2238. doi:10.1007/s11695-015-1710-y

Chahal D, Marquez V, Hussaini T, et al. End stage liver disease etiology & transplantation referral outcomes of major ethnic groups in British Columbia, Canada. *Medicine*. 2021;100(42). doi:10.1097/md.0000000000027436

Chinnaratha MA, Chelvaratnam U, Stuart KA, et al. Liver transplantation outcomes for Australian Aboriginal and Torres Strait islanders. *Liver Transplantation*. 2014;20(7):798-806. doi:10.1002/lt.23894

Clark S, Boyle L, Matthews P, Schweder P, Deng C, Campbell D. Development and validation of a multivariate prediction model of perioperative mortality in Neurosurgery. *Neurosurgery*. 2020;87(3). doi:10.1093/neuros/nyaa144

Ezomo OT, Sun D, Gronbeck C, Harrington MA, Halawi MJ. Where do we stand today on racial and ethnic health disparities? an analysis of primary total hip arthroplasty from a 2011–2017 national database. *Arthroplasty Today*. 2020;6(4):872-876. doi:10.1016/j.artd.2020.10.002

Fuchs HF, Broderick RC, Harnsberger CR, et al. Variation of outcome and charges in operative management for Diverticulitis. *Surgical Endoscopy*. 2014;29(11):3090-3096. doi:10.1007/s00464-014-4046-0

Hong Z, Wu J, Smart G, et al. Survival analysis of liver transplant patients in Canada 1997–2002. *Transplantation Proceedings*. 2006;38(9):2951-2956. doi:10.1016/j.transproceed.2006.08.180

Keel S, Xie J, Foreman J, Taylor HR, Dirani M. Population-based assessment of visual acuity outcomes following cataract surgery in Australia: The National Eye Health Survey. *British Journal of Ophthalmology*. 2018;102(10):1419-1424. doi:10.1136/bjophthalmol-2017-311257

Masoomi H, Kang CY, Chen A, et al. Predictive factors of in-hospital mortality in colon and rectal surgery. *Journal of the American College of Surgeons*. 2012;215(2):255-261. doi:10.1016/j.jamcollsurg.2012.04.019

Mordhorst TR, Jalali A, Nelson R, Brodke DS, Spina N, Spiker WR. Cost analysis of primary single-level lumbar discectomies using the value driven outcomes database in a large academic center. *The Spine Journal*. 2021;21(8):1309-1317. doi:10.1016/j.spinee.2021.03.017

Olson S, Law A. Meningiomas and the polynesian population. *ANZ Journal of Surgery*. 2005;75(8):705-709. doi:10.1111/j.1445-2197.2005.03499.x

Sequist TD, Narva AS, Stiles SK, Karp SK, Cass A, Ayanian JZ. Access to renal transplantation among American Indians and Hispanics. *American Journal of Kidney Diseases*. 2004;44(2):344-352. doi:10.1053/j.ajkd.2004.04.039

Shah BR, Frymire E, Jacklin K, Jones CR, Khan S, Slater M, Walker JD, Green ME. Peripheral arterial disease in Ontario First Nations people with diabetes: a longitudinal population-based cohort study. *Canadian Medical Association Open Access Journal.* 2019;7(4):E700-5.

Shively D, Makhani SS, Bouz A, Hernandez E, Chung-Bridges K. Racial Disparities in Survival Outcomes of Colorectal Cancer Patients After Surgical Resection. *Cureus.* 2022;14(2):e22064. doi: 10.7759/cureus.22064. PMID: 35295347; PMCID: PMC8916922.

Singleton, Neal, Emily Buddicom, Andrew Vane, and Vaughan Poutawera. Are there differences between Māori and non-Māori patients undergoing primary total hip and knee arthroplasty surgery in New Zealand? A registry-based cohort study. *New Zealand Medical Journal*. 2013;126(1379):23-30

Zafar S, Dun C, Srikumaran D, et al. Endophthalmitis rates among Medicare beneficiaries undergoing cataract surgery between 2011 and 2019. *Ophthalmology*. 2022;129(3):250-257. doi:10.1016/j.ophtha.2021.09.004

c. **Multiple Publications**

Alvord L, Rhoades D, Henderson W et al. Surgical Morbidity and Mortality among American Indian and Alaska Native Veterans: A Comparative Analysis. *Journal of the American College of Surgeons*. 2005;200(6):837-844. doi:10.1016/j.jamcollsurg.2005.01.015

Amirian SH. Predictors for Reoperation during 30-Day Period after Primary Bariatric Surgery (Doctoral dissertation, Rush University); 2019

Banham D, Roder D, Keefe D et al. Disparities in breast screening, stage at diagnosis, cancer treatment and the subsequent risk of cancer death: a retrospective, matched cohort of aboriginal and non-aboriginal women with breast cancer. *BMC Health Services Research*. 2019;19(1). doi:10.1186/s12913-019-4147-5

Barraclough K, Grace B, Lawton P, McDonald S. Residential Location and Kidney Transplant Outcomes in Indigenous Compared With Nonindigenous Australians. *Transplantation*. 2016;100(10):2168-2176. doi:10.1097/tp.0000000000001007

Cass A, Gillin AG, Horvath JS. End‐stage renal disease in Aboriginals in New South Wales: a very different picture to the Northern Territory. *Medical Journal of Australia*. 1999;171(8):407-410. doi:10.5694/j.1326-5377.1999.tb123718.x

Chan S, Pascoe EM, Clayton PA, McDonald SP, Lim WH, Sypek MP, Palmer SC, Isbel NM, Francis RS, Campbell SB, Hawley CM. Infection-related mortality in recipients of a kidney transplant in Australia and New Zealand. *Clinical Journal of the American Society of Nephrology*. 2019;14(10):1484-92.

Garrard E, McDonald S. Improving access to and outcomes of kidney transplantation for Aboriginal and Torres Strait Islander people in Australia: performance report. *Australian Indigenous Health Bulletin*. 2019;19(2).

Grace BS, Clayton PA, Cass A, McDonald SP. Transplantation rates for living-but not deceased-donor kidneys vary with socioeconomic status in Australia. *Kidney International*. 2013;83(1):138-45.

Howson P, Irish AB, D’Orsogna L, Chakera A, Swaminathan R, Perry G, De Santis D, Tolentino R, Wong G, Lim WH. Allograft and patient outcomes between indigenous and nonindigenous kidney transplant recipients. *Transplantation*. 2020;104(4):847-55.

LaPar D, Bhamidipati C, Harris D et al. Gender, Race, and Socioeconomic Status Affects Outcomes After Lung Cancer Resections in the United States. *Annals Thoracic Surgery*. 2011;92(2):434-439. doi:10.1016/j.athoracsur.2011.04.048

Lovrics O, Doumouras AG, Gmora S, Anvari M, Hong D. Metabolic outcomes after bariatric surgery for Indigenous patients in Ontario. *Surgery for Obesity and Related Diseases*. 2019;15(8):1340-7.

McDonald SP, Russ G. Current incidence, treatment patterns and outcome of end-stage renal disease among indigenous groups in Australia and New Zealand. *Nephrology*. 2003;8(1):42-48. doi:10.1046/j.1440-1797.2003.00131.x

McDonald SP, Russ GR. Survival of recipients of cadaveric kidney transplants compared with those receiving dialysis treatment in Australia and New Zealand, 1991–2001. *Nephrology Dialysis Transplantation*. 2002;17(12):2212-9

McDonald S. Indigenous transplant outcomes in Australia: What the ANZDATA Registry tells us. *Nephrology*. 2004;9(s4):S138-S143. doi:10.1111/j.1440-1797.2004.00350.x

Nguyen N, Masoomi H, Laugenour K et al. Predictive factors of mortality in bariatric surgery: Data from the Nationwide Inpatient Sample. *Surgery*. 2011;150(2):347-351. doi:10.1016/j.surg.2011.05.020

Rogers N, Lawton P, Jose M. Kidney Transplant Outcomes in the Indigenous Population in the Northern Territory of Australia. *Transplantation*. 2006;82(7):882-886. doi:10.1097/01.tp.0000232439.88527.49

Russell E, Walsh W, Reid C et al. Outcomes after mitral valve surgery for rheumatic heart disease. *Heart Asia*. 2017;9(2):e010916. doi:10.1136/heartasia-2017-010916

Wang T, Wei D, Evans T, Haydock D, Ramanathan T. Ethnic Comparisons of Type A Aortic Dissection Presentation and Outcomes. *Heart, Lung and Circulation*. 2019;28:S59-S60. doi:10.1016/j.hlc.2019.05.153

Weber C, Rush D, Jeffery J, Cheang M, Karpinski M. Kidney Transplantation Outcomes in Canadian Aboriginals. *American Journal of Transplantation*. 2006;6(8):1875-1881. doi:10.1111/j.1600-6143.2006.01409.x

Wiemers P, Marney L, White N et al. Comorbidities and Ventricular Dysfunction Drive Excess Mid-Term Morbidity in an Indigenous Australian Coronary Revascularisation Cohort. *Heart, Lung and Circulation*. 2019;28(6):874-883. doi:10.1016/j.hlc.2018.04.285

Wiemers P, Muller R, Brandon M et al. Cardiac Surgery in Indigenous Australians—How Wide is ‘The Gap’?. *Heart, Lung and Circulation*. 2011;20(4):251-252. doi:10.1016/j.hlc.2010.11.010
