## Supplementary material for "Post-operative morbidity and mortality in Indigenous Peoples: A scoping review and meta-analysis": S1 Table

**S1 Table: Included study descriptions and summary of findings**

| **First author, year** | **Study Design** | **Age group; participants (n)** | **Surgical discipline; procedure(s)** | **Outcome(s)** | **Overall difference in M&M for IN?**** | **Overall Quality Assessment** |
| --- | --- | --- | --- | --- | --- | --- |
| ***North America*** | | | | | | |
| **Canada** | | | | | | |
| Chahal, 2021 | Retrospective cohort | NR*; First Nations n=235; Caucasian n=2725 | General Surgery (HPB / Transplant); liver transplant | **Immunologic Complications**: acute rejection; chronic rejection; graft survival | No difference | Poor |
| Cohen, 1989 | Retrospective cohort | NR*; Native American n=1327; Comparator Other n=36056 | General Surgery; Cholecystectomy | **Readmission:** all post-op readmissions; readmission to hospital after cholecystectomy | Increase | Poor |
| Goulet, 2006 | Retrospective cohort | Adult (>18); Aboriginal n=108 procedures; Non-Aboriginaln=720 procedures | Vascular Surgery; PVD revascularization (bypass) | **<30-day Mortality**; **Surgical Infections:** wound infection at 30 days; **CV Complications:** loss of limb at 5yrs; MI at 30 days; **H&T Complications:** graft occlusion at 5 yrs | Increase | Good |
| Hong 2006 | Retrospective Cross-sectional | NR; Total n=1122*; Aboriginal n = NA; Caucasian n =NA | General Surgery (HPB / Transplant); liver transplant | **>30-Day Mortality:** 1yr post-transplant survival | Increase | Poor |
| Meatherall, 2005 | Cross-sectional | Adult (>18); Aboriginal n=21; Non-Aboriginal n=23 | Orthopedic Surgery; lower extremity amputation | **Surgical Infection:** wound at amputation site; **Procedural Complications:** pain | Increase | Poor |
| Shah, 2019 | Retrospective cohort | Adult (>18); First Nations n =36584; Others n =2045015 | Vascular Surgery; Orthopedic Surgery: below the knee amputation | **>30-Day Mortality: Survival:** mortality; survival | Increase | Good |
| Sood, 2013 | Retrospective cohort | All (>15); Aboriginal n = 574; Non-Aboriginal n = 11596 | Cardiac Surgery: CABG; valve replacement | **Overall Morbidity:** post-op infections; major adverse event (ACS, stroke, mortality, need for dialysis) **Pulmonary Complications:** pneumonia; **<30 day Mortality:** post-op mortality | No difference | Good |
| Storsley, 2010 | Retrospective cohort | Aboriginal n = 38; White n = 76 | Urology/General Surgery: renal transplant | **CV Complications:** post-transplant hypertension; **Immunologic Complications:** post-op development of diabetes; **GU Complications:** proteinuria | Increase | Poor |
| Zhang, 2011 | Retrospective Cross-sectional | All (>16); FN n=20; Non FNs n = 129 | General Surgery (HPB / Transplant); liver transplant | **>30-Day Mortality:** rejection-free survival; overall survival | No difference | Poor |
| **USA** | | | | | | |
| Al-Qurayshi, 2016 | Retrospective cross-sectional | Adult (>18); NA n=4287; White n=13247 | General Surgery (Endocrine)/ENT; thyroidectomy | **Overall Morbidity:** post-op complications | No difference | Good |
| Alvord, 2009 | Cross-sectional | Adult (>18); AI/NA n=2155; Caucasian n=2264 | General Surgery/Orthopedic Surgery/Thoracic Surgery/Urology/Vascular Surgery; inguinal hernia repair, colectomy, laparoscopic cholecystectomy, arthroplasty, laminectomy, pulmonary lobectomy, TURP, radical prostatectomy, endarterectomy, amputations, revascularization | **Surgical Infections:** wound infection; deep wound infection; dehiscence; **CV Complications:** cardiac arrest; pulmonary edema; MI; **H&T Complications:** CVA; PE; DVT/SVT; >4u RBC transfusion; **Immunologic Complications:** graft/prosthesis rejection; **GU Complications:** UTI; progressive RI, ARF; **Procedural Complications:** ileus; peripheral nerve injury; **Pulmonary Complications:** pneumonia; Intubation >48h; **Systemic Infections:** sepsis | Increase | Good |
| Amirian, 2020 | Cross-sectional | Adult (>18); AI/NA n=377, NH/PI n=340; Caucasian n=85054 | General Surgery (Bariatric); laparoscopic sleeve gastrectomy, Roux-en-Y | **Systemic Infections:** post-op sepsis; post-op septic shock, C. difficile infection; **Reoperation:** at least 1 reoperation ≤ 30 days; **Readmission:** at least 1 readmission ≤ 30 days; **LOS; GU Complications:** post-op UTI; ARF; progressive RI; **Surgical Infections:** post-op superficial SSI; deep incisional SSI; organ/space SSI; wound interruption; **Pulmonary Complications:** on ventilator >48hrs; post-op pneumonia; unplanned intubation; **H&T Complications:** DVT; stroke/CVA; PE; **CV Complications:** intra-op or post-op cardiac arrest requiring CPR; intra-op or post-op MI; **GU Complications:** post-op UTI; ARF; progressive RI; **Procedural Complications:** coma >24hr; unplanned ICU admission; operative drain still present at 30 days; incisional hernia | Increase | Good |
| Anderson, 2018 | Retrospective cohort study | Adult (>18); AI n=74; Non-AI n=1236 | Cardiac Surgery; CABG | **<30-Day Mortality:** operative mortality; **LOS:** LOS >14; **Overall Morbidity:** major morbidity; **Reoperation**; **H&T Complications:** permanent stroke | No difference | Poor |
| Armenia, 2017 | Cross-sectional | Adult (>18); Native American n = 1438; White n=283376 | General Surgery (Acute Care); multiple emergency surgeries (non-elective) | **<30-Day Mortality:** in-hospital mortality | Increase | Good |
| Betancourt-Garcia, 2019 | Cross-sectional | Adult (>18); AI/AN; Pop n=20232; White n=2433675 | Cardiac Surgery; multiple (no trauma, transplant, ASA >6, or minor cases) | **LOS; <30-Day Mortality**; **Systemic Infections:** sepsis; septic shock; **Pulmonary Complications:** ventilation >48 hours; SSI (deep, organ space, wound disruption) | NR | Good |
| Boyd, 2021 | Retrospective cohort | Adult (>18); AI/AN p n=85; White n=16093 | Obstetrics & Gynecology; vaginal colpopexy, abdominal sacrocolpopexy | **Procedural Complications:** Clavien-Dindo complication grade II; **GU Complications:** UTI; **H&T Complications:** blood transfusion | No difference | Good |
| Broderick,  2015 | Retrospective cross-sectional | Adult (>18); Native American n=849; White n=124086 | General Surgery (Bariatrics) | **<30-Day Mortality:** in-hospital mortality | No difference | Good |
| Brown, 2020 | Retrospective cohort | Adult (>18); AI/AN Pop n=289; NH n=257; Caucasian n=34254 | Obstetrics & Gynecology; laparoscopic hysterectomy | **Surgical Infections:** deep and organ space SSI | No difference | Good |
| Causey, 2013 | Cross-sectional | Adult (>18);AI/AN n=968; White n=60093 | General surgery (Acute Care); relief of bowel obstruction, perforated hollow viscus repair, colectomy, incarcerated hernia repair, cholecystectomy, appendectomy | **Overall Morbidity:** 30-day complication rate (from NSQIP); <**30-Day Mortality** | Increase | Good |
| Changoor, 2015 | Cross-sectional | Adult (>18); AI/AN n=1968; NH/PI n = 490; White n=145610 | General Surgery; laparoscopic cholecystectomy, laparoscopic appendectomy, open hernia repair | **Overall Morbidity:** major complications; minor complications; **Surgical Infection:** wound infections; <**30-Day Mortality**; **Reoperation:** return to OR | NR | Poor |
| Chen, 2020 | Retrospective cross-sectional | Adult (>18); Native American n=3296; White n=610065 | General Surgery; colectomy, gastrectomy, hepatectomy, pancreatectomy | **H&T Complications:** DVT; PE; post-op hemorrhage; **LOS:** LOS >7 days | Increase | Good |
| Chertack, 2021 | Cross-sectional | Adult (>18); Native American n=1157; NHW n=190578 | Urology; urologic surgeries | **Overall Morbidity:** any complication; minor complications (superficial SSI, pneumonia, PE, UTI, transfusion, DVT); major complication (deep SSI, organ space infection, wound dehiscence, reintubation, ventilation,RI, ARF, stroke, cardiac arrest, MI, sepsis, septic shock); **>30-Day Mortality** | No difference | Good |
| Ezomo, 2020 | Retrospective cross-sectional cohort | Adult (>18); AI/AN n=594; NH/PI n = 417; NHW n=116129 | Orthopedic Surgery; total hip arthroplasty | **<30-Day Mortality**; **Readmission**: 30-day readmission; **Reoperation**; **LOS**; **Overall Morbidity:** any complication | Increase | Good |
| Fuchs,  2015 | Retrospective cross-sectional cohort | Adult (>18); Native American n=383; White n=96858 | General Surgery (Colorectal); colon and rectal resections | **>30-Day Mortality:** mortality | No significant difference | Good |
| Holleran 2021 | Retrospective cohort | Adult (>18); Native American n=290; Caucasian n=27907 | General Surgery (Colorectal); colon and rectal resections | **<30-Day Mortality**; **LOS; Overall Morbidity**; **Surgical Infections:** wound complications; **Systemic Infections:** sepsis | Decrease (LOS) | Good |
| Hsieh, 2020 | Retrospective cross-sectional cohort | Adult (>18); Native American n=837; Caucasian n=55098 | Obstetrics & Gynecology; cesarean section | **Surgical Infections:** SSI; **H&T Complications**: blood transfusion;amniotic embolism;PPH;DIC;hematoma; **CV Complications:** cardiac arrest;vfib;pulmonary edema; **LOS**; **Procedural Complications:** severe anesthetic complications | Increase | Poor |
| Ivan,  2021 | Retrospective cohort | Adult (>18); Native American n=12; Non-Native American n=62 | Vascular Surgery: orbital atherectomy | **CV Complications:** MACE events (1yr postop); amputations; <**30-Day Mortality; Reoperation:** reintervention rate | Increase | Poor |
| Kamaraju, 2021 | Retrospective cohort | Adult (>18);Native American/AN n=1675; NH/PI n = 1243; White n=159459 | Orthopedic Surgery; arthroplasty (unicondylar versus total knee) | **LOS; Readmission:** 30-day readmission; <**30-Day Mortality**; **Overall Morbidity:** UKA overall complications; TKA overall Complications | No difference | Good |
| Kasiske, 1998 | Retrospective cohort | NR*; AI/AN n=68; Caucasian=1253 | Urology/General Surgery (Transplant); renal transplant | **>30-Day Mortality; Immunologic Complications:** graft failure; acute rejection; DGF | No difference | Poor |
| Kwan, 2018 | Retrospective cohort | NR*; AI n=1908; NH/PI n=843; White n=110524 | Urology/General Surgery (Transplant); renal transplant | **Immunologic Complications:** DGF; acute rejection; graft failure; **GU Complications:** proteinurea | Increase | Poor |
| Mahdi, 2014 | Retrospective cross-sectional cohort | All ages; AI/AN n=621; NHW n=97763 | Obstetrics & Gynecology; endometrial cancer surgeries (including hysterectomy, LND, radical hysterectomy) | **>30-Day Mortality:** overall survival; cancer-specific survival | Increase | Poor |
| Markin, 2013 | Retrospective cohort | NR*; AI n=3052; NHW n=737826 | General Surgery (Surgical Oncology); surgery for oncologic tumors | **<30-Day Mortality:** in-hospital mortality; **LOS:** prolonged LOS | No difference | Poor |
| Martin, 2017 | Retrospective cohort | NR*; AI n=7; White n=1054 | Neurosurgery; spine surgeries | **Readmission** | No difference | Poor |
| Mordhorst, 2021 | Retrospective cohort | Adult; Native American n=4; White n=564 | Neurosurgery; lumbar discectomies | **LOS** | No difference | Good |
| Masoomi,  2012 | Retrospective cross-sectional cohort | Adult (>18); Native American n=4879; Whiten=771878 | General Surgery (Colorectal); right colectomy, transverse colectomy, left colectomy, sigmoidectomy, total colectomy, proctectomy, other unspecified partial/segmental intestinal resections/excisions | **<30-Day Mortality:** in-hospital mortality | No difference | Good |
| Nallamothu, 2001 | Retrospective cross-sectional cohort | Adult (>18); Native American n=155; White n=16077 | Cardiac Surgery; CABG | **LOS:** **<30-Day Mortality:** In-hospital death | Increase | Poor |
| Roche, 2019 | Retrospective cross-sectional cohort | Adult (>18); Native American =261; White n=98257 | Orthopedic Surgery; arthroplasty | **Reoperation:** revision rate for TKA | No difference | Good |
| Sanford, 2019 | Cross-sectional | Adult (>18); Native American n=175; Caucasian n=4106 | Neurosurgery; spine surgeries (cervical, lumbar fusion, decompression laminectomy) | **Surgical Infections:** superficial SSI; organ space SSI; dehiscence; **CV Complications:** cardiac arrest; **Pulmonary Complications:** pneumonia; reintubation; **H&T Complications:** PE; DVT; **GU Complications:** UTI; **Reoperation:** return to OR; **Readmission:** 30-day readmission | Increase | Good |
| Seipp, 2021 | Case-Control | NR*; Indigenous n=165; White n=165 | Urology/General Surgery (Transplant); renal transplant | **Surgical Infections:** post-transplant infection; **Immunological complications:** acute rejection; DGF; **Readmission:** rehospitalization rate after transplant; **>30- Day Mortality**: all-cause mortality | No difference | Good |
| Sequist, 2004 | Retrospective cohort | NR*;AI=477; Whiten=470 | Urology/General Surgery (Transplant); renal transplant | **Immunological Complications:** graft failure | No significant difference | Good |
| Shively, 2021 | Retrospective cross-sectional cohort | Adult; AI n=903; White n=81019 | General Surgery (Colorectal / Surgical Oncology)); colorectal cancer resections | **>30-Day Mortality:** median survival; 1, 5yr survival | No difference | Good |
| Simianu, 2016 | Prospective cohort | Adult (>18); AI/AN n=156; NHWn=6030 | General Surgery (Surgical Oncology); surgery for oncologic tumors | **>30-Day Mortality; Overall Morbidity:** post-op complications; **LOS** | Increase | Poor |
| Tan, 2019 | Retrospective cohort | Adult (>18); Native American n=1631; White n=98361 | Orthopedic Surgery; amputation +/- revascularization | **LOS:** LOS post-minor amputation | Increase | Poor |
| Wiley, 2021 | Retrospective cohort | Adult (>18);Northern Great Plains Native American n=117; White n=505 | Urology/General Surgery (Transplant); renal transplant | **Immunological Complications:** rejection events <1 yr post-transplant; all cause graft failure at 1 mo, 1, 3, 5, 10yrs **Systemic Infections:** CMV positive; **>30-Day Mortality:** survival at 1, 3, 5, 10yrs; **GU Complications:** eGFR at 1 mo, 1, 3yrs | No difference | Good |
| Yang, 2020 | Retrospective cross-sectional | Adult; Native American n=1618; White n=42945 | Orthopedic Surgery; total hip arthroplasty | **Overall Morbidity:** prosthesis related complication (mechanical loosening, dislocation, periprosthetic joint infection, periprosthetic fracture, other) | Increase | Poor |
| Yang 2021 | Retrospective cross-sectional | Adult; Native American n=5,900; Whiten=1,023,445 | Orthopedic Surgery; total knee arthroplasty | **Overall Morbidity:** prosthesis related complication (mechanical loosening, dislocation, periprosthetic joint infection, periprosthetic fracture, other) | No difference | Poor |
| Zafar, 2021 | Retrospective cross-sectional | Adult; Native American n=71,082; White n=12,645,241 | Ophthalmology; cataract surgery | **Surgical Infections:**endophthalmitis rates | Increase | Good |
| Zhang, 2016 | Retrospective cross-sectional cohort | Adult; Native American n=2,792; White n=477,534 | Orthopedic Surgery; arthroplasty | **<30-Day Mortality:** in-hospital mortality, **LOS; Overall Morbidity:** in-hospital complications | Increase | Poor |
| ***Oceania*** | | | | | | |
| **Australia** | | | | | | |
| Alizzi, 2010 | Retrospective cohort | Any; Indigenous n=423; Non-Indigenous n=412 | Cardiac Surgery; CABG +/- valve replacement | **<30-Day Mortality:** operative mortality; 30-Day Mortality; **H&T Complications:** stroke; **Pulmonary Complications:** intubation >48h; **GU Complications:** post-op renal failure requiring dialysis | Increase | Poor |
| Banham, 2019 | Case-control | NR*; Aboriginal n=777; Non-Aboriginal n=777 | General Surgery (Surgical Oncology); surgery for oncologic tumors | **>30-Day Mortality:** cancer death post-op; cancer death after surgery, chemotherapy and radiotherapy | Increase | Good |
| Boan, 2017 | Case-control | Adult (>18); ATSI n=57; Non-ATSI n=84 | Urology/General Surgery (Transplant); renal transplant | **Immunologic Complications:** acute rejection; graft loss; **Surgical Infection:** readmission due to infection; **>30-Day Mortality:** 5 yr post-transplant survival | Increase | Poor |
| Chinnaratha, 2014 | Retrospective cohort | All ages; ATSI n=45; Non-ATSI n=3448 | General Surgery (HPB / Transplant); liver transplant | **>30-Day Mortality:** median overall survival | No difference | Poor |
| Elahi, 2008 | Retrospective cohort | Adult (>18); Aboriginal n=20; British Caucasian n=493 | Cardiac Surgery; CABG | **<30-Day Mortality; >30-Day Mortality:** 6 mo mortality; **LOS:** post-op and ICU LOS**; H&T Complications:** CVA;  **Pulmonary Complications**: pulmonary complications; mechanical ventilation >24h | Increase | Good |
| Henman, 2012 | Retrospective and prospective cohorts | NR*; Indigenous n=84; Non-Indigenous n=123 | Obstetrics & Gynecology; cesarean section | **Surgical Infections:** SSI | Increase | Poor |
| Keel, 2018 | Retrospective cross-sectional | Adult (>18);Indigenous n=223; Non-Indigenous n=1110 | Ophthalmology; cataract surgery | **Overall Morbidity:** surgery complication | Increase | Poor |
| Keenan, 2019 | Retrospective cross-sectional cohort | NR*; Indigenous n=82; Non-Indigenous n=154 | Cardiac Surgery; re-do valve repair | **LOS:** median hospital stay; <**30-Day Mortality**; **Reoperation:** Return to OR; **Pulmonary Complications:** prolonged ventilation >48 hrs; pneumonia; **H&T Complications:** stroke; blood transfusion; **GU Complications:** AKI including those needing dialysis; **Overall Morbidity:** combined morbidity (stroke, MI, post-op dialysis, mechanical ventilation >48hr, deep sternal wound infection or reoperation); **Surgical Infections:** sternal wound infection | No difference | Poor |
| Lehman, 2009 | Prospective cohort | NR*; Indigenous =283; Non-Indigenous n=2352 | Cardiac Surgery; valve surgeries +/- CABG | **H&T complications:** postop stroke;**GU Complications:** postop renal failure; **30-Day Mortality:** operative mortality | Increase | Good |
| Majoni, 2019 | Retrospective cross-sectional | NR*; Indigenous n=343; Non-Indigenous n=6207 | Urology/General Surgery (Transplant); renal transplant | **Immunologic Complications:** rejection; DGF | Increase | Good |
| O'Brien, 2018 | Retrospective cohort | Adult (>18); Indigenous 778; Non-Indigenous n=36124 | Cardiac Surgery; CABG | **Surgical Infections:** deep sternal wound infection; **>30-Day Mortality:** mortality (total); cardiac death; <**30-Day Mortality; H&T Complications:** bleeding complication; **Readmission:** 30-day hospital readmission; | No significant difference | Poor |
| O'Rourke, 2013 | Retrospective cohort | Adult (>18); Indigenous n = 74; Non-Indigenous n = 69 | Orthopedic Surgery: below the knee amputation | **LOS** | Increase | Poor |
| Page, 2021 | Retrospective cohort | Adult (>18); Indigenous n=42; Non-Indigenous n=30 | Cardiac Surgery; Isolated CABG, CABG + valve surgery, or valve surgery (with or without aortic root) | **Reoperation:** return to OR; <**30-Day Mortality:** early mortality; in-hospital mortality; **>30-Day Mortality:** late mortality ; **Surgical Infections:** deep sternal wound infection | No difference | Good |
| Prabhu, 2013 | Prospective cohort | Adult (>18); Indigenous n=297; Non-Indigenous n=2451 | Cardiac Surgery; CABG | **LOS; <30-Day Mortality;** **>30-Day Mortality:** all-cause mortality **Reoperation**; **Overall** **Morbidity:** post-op complications (mortality, stroke, CVA, renal dialysis, ventilation >24hrs, sternal wound infection, reoperation, MI); **Surgical Infections:** sternal wound infection | Increase | Good |
| Rogers, 2011 | Retrospective cohort | NR*; ATSI n=108; Non-Indigenous n=69 | Urology/General Surgery (Transplant); renal transplant | **Immunologic Complications:** all cause allograft failure and allograft survival; any biopsy-proven rejection; acute rejection | Increase | Poor |
| Russell, 2015 | Retrospective cross-sectional | NR; Indigenous n=174; Non-Indigenous n=1,210 | Cardiac surgery; valve surgery for rheumatic heart disease | **<30-Day Mortality**; **>30-Day Mortality:** long-term mortality, **LOS:** post-op LOS; **GU Complications:** post-op AKI; **CV Complications:** post-op AFib, | No difference | Poor |
| Singh, 2021 | Retrospective cohort | NR*; ATSI n=70; Non-Indigenous n=304 | Orthopedic Surgery; amputation | **>30-Day Mortality:** all-cause mortality; **Overall Morbidity:** contralateral amputation | Increase | Poor |
| Swift,  2021 | Retrospective cross-sectional | All; Indigenous n=661; Non-Indigenous n=1,073 | General Surgery; appendectomy | **LOS:** median LOS; extended LOS | Increase | Poor |
| Treacy, 2015 | Retrospective cross-sectional | NR; Aboriginal n=72; Non-Aboriginal n=118 | All specialties, multiple surgeries | **Overall Morbidity; Reoperation:** unplanned return to OR | No difference | Good |
| Treacy, 2016 | Prospective cohort | Adult (>18); metro Indigenous n=33; metro non-Indigenous n=433 | General Surgery (Bariatric); gastric banding | **Reoperation**: reoperation; band removal/revision; **Surgical Infections:** erosion/infection; **Procedural Complications:** other post-op complications (slip, leak, band intolerance, port problem) | No difference | Poor |
| Weir, 2016 | Retrospective cross-sectional | Adult; Aboriginal n=212; Non-Aboriginal n=23,292 | General Surgery (Colorectal / Surgical Oncology);colorectal cancer resections) | **<30-Day Mortality:** post-op mortality, 30-day post-op mortality, **Overall Morbidity:** surgical complications | Increase | Good |
| **New Zealand** | | | | | | |
| Chiang, 2018 | Retrospective cohort | Adult (>18); Māori  n=95; European n=900 | Vascular Surgery; AAA Repair | **LOS**; <**30-Day Mortality**; **>30-Day Mortality:** 1, 5, 10yr mortality | Increase | Poor |
| Clark, 2020 | Retrospective cohort | Adult (>18); Māori n=1831; European n=10075 | Neurosurgery; neurosurgical operations | **<30-Day Mortality**; **>30-Day Mortality:** 1, 2yr mortality | NR | Good |
| D'Costa, 2001 | Retrospective cohort | All ages (12-60); Māori/Polynesian n=23; NZ European n=79 | Cardiac Surgery; heart transplant | **>30-Day Mortality:** 1, 5yr survival; **Immunologic Complications:** transplant rejection | No difference | Poor |
| Gurney,  2018 | Retrospective cross-sectional | Adult; Māori n=1,297; PI n=551; European/other n=4339 | Orthopedic Surgery; Low limb amputation | **30 day mortality; >30 day mortality:** 90-day mortality | Increase | Good |
| Khashram, 2017 | Retrospective cohort | Adult (>18); NZ Māori n=421; pI n = 97; NZ European n=5654 | Vascular Surgery; AAA Repair | **LOS**, <**30-Day Mortality**; **>30-Day Mortality:** overall survival | Increase | Good |
| Kwon, 2017 | Retrospective cohort | NR*; Māori n=189; PI n=136; European n=210 | General Surgery (Endocrine) and ENT; thyroid surgery | **H&T Complications:** hemorrhage; **LOS**; **Readmission:** readmission rate | Increase | Poor |
| Lao, 2019 | Retrospective cross-sectional cohort | All (14-99); Māori n=5988; European n=55545 | Orthopedic Surgery; arthroplasty (primary hip and knee) | **<30-Day Mortality**; **>30-Day Mortality:** 90-day, 1yr mortality; standardized mortality ratio of primary hip replacement | NR | Good |
| Olson, 2005 | Retrospective cohort | Adult (>18); Māori & PI=151; NZ European n=130 | Neurosurgery; tumor resection (meningioma) | **<30-Day Mortality**: death by 3-week follow-up;  **Procedural Complications**: neurological deficit | No difference | Poor |
| Pai, 2010 | Prospective cohort | Adult (>18); Māori n=38; Caucasian n=52 | Orthopedic Surgery; arthroplasty | **Surgical Infections:** superficial/deep SSI; **LOS**; **H&T Complications:** hematoma/wound ooze (bleeding); VTE | No difference | Poor |
| Singleton, 2013 | Prospective cross-sectional cohort | Adult (>18); Māori n=329; Non-Māori n=2061 | Orthopedic Surgery; total hip and knee arthroplasty | **Overall Morbidity:** WOMAC Total, SF12 Mental Health, WOMAC Stiffness, and SF12 Physical Health scores 1, 5 yrs post-op | Increase | Poor |
| Stewart, 2014 | Retrospective cohort | NR; Māori n=47; Non-Māori n=206 | Cardiac Surgery; heart transplant | **>30-Day Mortality:** 5, 10yr mortality; **LOS:** post-transplant LOS | Increase | Poor |
| Swart, 2013 | Case-Control | Adult; Māori n=70; Non-Māori n=74 | General Surgery (Colorectal / Surgical Oncology); rectal cancer resections | **Pulmonary Complications:** pneumonia; **Systemic Infections:** sepsis; **Reoperation:** reoperation due to anastomotic leakage, bleeding, intra-abdominal abscess; **Overall Morbidity:** organ failure (cardiac, respiratory, renal) | No difference | Good |
| Wu, 2020 | Retrospective cohort | NR; Māori n=37; non-Māori n=56 | General Surgery (Endocrine) and ENT; thyroid surgery | **Procedural Complications:** RLN injury; procedural complications | Increase | Poor |
| Wang, 2013 | Retrospective cohort | Adult; Māori n=82; European descent n=444 | Cardiac Surgery; CABG | **<30-Day Mortality**, **Overall Morbidity:** composite surgical morbidity, **Readmission:** readmission to hospital within 30 days; **Reoperation;**, **Surgical Infections:** deep sternal wound infection | Increase | Poor |
| Wang, 2020 | Prospective cohort | Adult; Māori n=45; PI n=91; Other n=191 | Cardiac Surgery/Vascular Surgery; aortic dissection repair | **>30-Day Mortality:** late mortality; <**30-Day Mortality:** operative mortality; **Overall Morbidity:** composite morbidity (stroke; AKI; ventilation >24 hrs; deep sternal wound infection; return to OR); **LOS**; **Surgical Infections:** sternal wound infection; **Reoperation**; **H&T Complications:** stroke; **CV Complications:** pacemaker insertion; **Pulmonary Complications:** ventilation >24 hrs; **GU Complications:** AKI | Increase | Good |

NR = Not recorded in study

Participant populations are listed as reported in individual studies

*population of interest and comparator population not differentiated

**Increase refers to at least one outcome of interest being reported as significantly worse for Indigenous populations versus non-Indigenous comparators; if both decrease and increase in outcomes reported, the specific outcome is reported in brackets; no significant difference denotes no outcome reported as better or worse for morbidity and mortality in Indigenous patients; NR reflects lack of significance (e.g. p-value) provided for outcomes of interest in the study

**Abbreviations**

AAA	Abdominal Aortic Aneurysm

ACS	Acute Coronary Syndrome

ARF:	Acute Renal Failure

CABG	Coronary Artery Bypass Graft

CHD	Congenital Heart Disease

CV	Cardiovascular

CVA	Cerebrovascular Accident / Stroke

DGF	Delayed Graft Function

DVT	Deep Vein Thrombosis

GU	Genitourinary

HLHS	Hypoplastic Left Heart Syndrome

HPB	Hepatobiliary-Pancreatic Surgery

H&T	Hematologic and Thromboembolic

I&D	Incision & Drainage

LND	Lymph Node Dissection

LOS	Length of Stay

M&M	Morbidity & Mortality

NA	Not Applicable

NSQIP	National Surgical Quality Improvement Program

PE	Pulmonary Embolism

RI	Renal Insufficiency

PVD	Peripheral Vascular Disease

SSI	Surgical Site Infection

SVT	Superficial Vein Thrombosis

TAVI	Transcatheter Aortic Valve Implantation

TURP	Transurethral Resection of the Prostate

UTI	Urinary Tract Infection

**Population Abbreviations**

AI	American Indian

AN	Alaskan Native

ATSI	Aboriginal and Torres Strait Islander

NH	Native Hawaiian

NHW	Non-Hispanic White

NZ	New Zealand

PI	Pacific Islander
