## Supplementary material for "Post-operative morbidity and mortality in Indigenous Peoples: A scoping review and meta-analysis": S2 Table

**S2 Table: Operations included in studies**

| **Surgical Specialty and Associated Procedures** | **No. of studies reporting*** |
| --- | --- |
| **General Surgery** |  |
| Appendectomy; laparoscopic appendectomy | 2 |
| Gastric banding; gastric bypass; sleeve gastrectomy; gastric banding; Roux-en-Y | 3 |
| Colectomy; right colectomy; transverse colectomy; left colectomy; sigmoidectomy; total colectomy; proctectomy; other unspecified partial/segmental intestinal resections/excisions; colorectal cancer resections; colon and rectal resections | 5 |
| Pancreatectomy | 1 |
| Hepatectomy | 1 |
| Liver transplant | 4 |
| Inguinal hernia repair; open hernia repair | 2 |
| Cholecystectomy; laparoscopic cholecystectomy | 2 |
| Gastrectomy | 1 |
| Colorectal cancer resection; rectal cancer resection; oncologic tumor resection | 6 |
| Multiple emergency surgeries | 2 |
| **ENT** |  |
| Thyroidectomy; thyroid surgery | 3 |
| **Vascular** |  |
| AAA repair | 2 |
| PVD revascularization (bypass) | 1 |
| Orbital atherectomy | 1 |
| Endarterectomy | 1 |
| **Cardiac Surgery** |  |
| CABG | 10 |
| Valvular replacement; re-do valve repair | 5 |
| Heart transplant | 3 |
| Type A aortic dissection repair (including aortic and mitral valves repairs, CABG, and/or partial arch repair) | 1 |
| Multiple surgeries (unspecified) | 1 |
| **Thoracic** |  |
| Pulmonary lobectomy | 1 |
| Lung tumor resection | 1 |
| **Orthopedic Surgery** |  |
| Arthroplasty (primary hip/knee) | 9 |
| Amputation; amputation and revascularization; lower extremity amputation; below the knee amputation | 6 |
| Laminectomy | 1 |
| **Urology** |  |
| Kidney transplant | 9 |
| Urologic surgeries (unspecified) | 1 |
| TURP | 1 |
| Radical prostatectomy | 1 |
| **Obstetrics and Gynecology** |  |
| Cesarean section | 2 |
| Endometrial cancer surgery (including hysterectomy, LND, radical hysterectomy) | 1 |
| Laparoscopic hysterectomy | 1 |
| Vaginal colpopexy; abdominal sacrocolpopexy | 1 |
| **Ophthalmology** |  |
| Orbital atherectomy | 1 |
| Cataract surgery | 2 |
| **Neurosurgery** |  |
| Spine surgeries (unspecified) | 2 |
| Lumbar discectomies | 1 |
| Neurosurgical operations (unspecified) | 1 |
| Tumor resection | 1 |
| **All Specialties** |  |
| All surgeries (unspecified) | 1 |

*****One study can contribute to more than one category and/or specialty

CABG: Coronary Artery Bypass Grafting; ENT: Ears, Nose, Throat; LND: Lymph Node Dissection; OBGYN: Obstetrics and Gynecology; TURP: Transurethral Resection of the Prostate
