## Supplementary material for "Post-operative morbidity and mortality in Indigenous Peoples: A scoping review and meta-analysis": S5 Appendix

**S5 Appendix: Funnel plots for publication bias assessment**

| 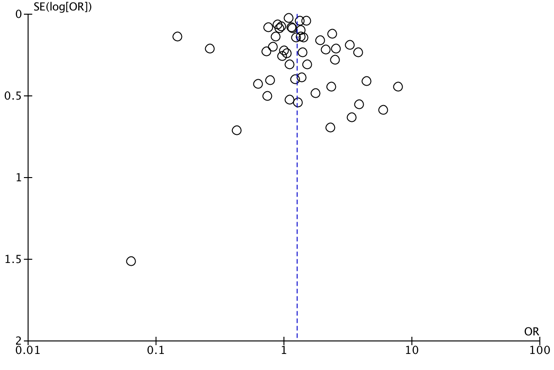 | 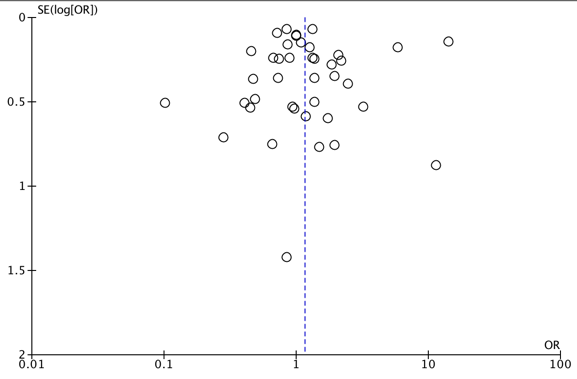 |
| --- | --- |
| Overall Morbidity | Overall Mortality |
| 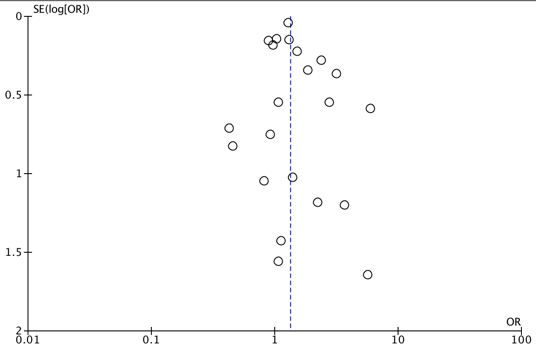 | 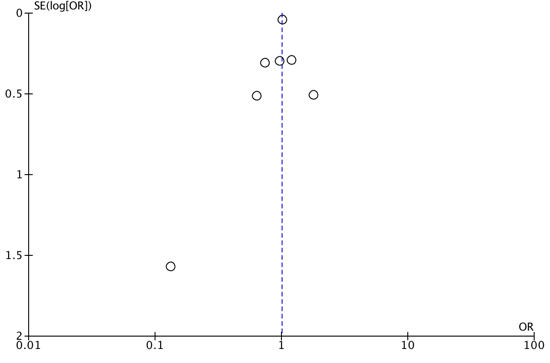 |
| Surgical Infections | Systemic Infections |
| 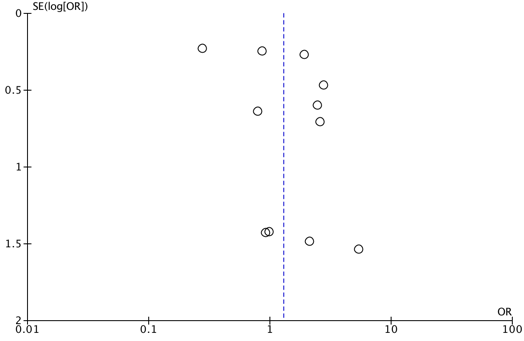 | 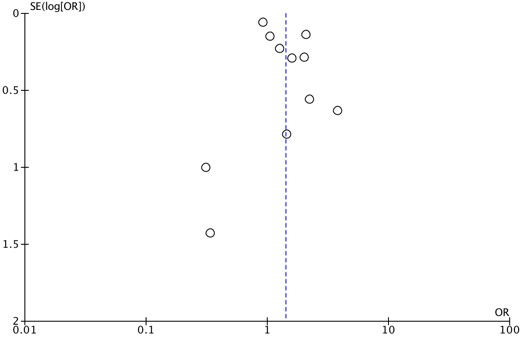 |
| Cardiovascular Complications | Pulmonary Complications |
| 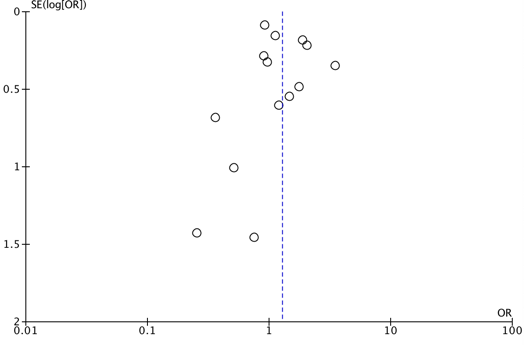 | 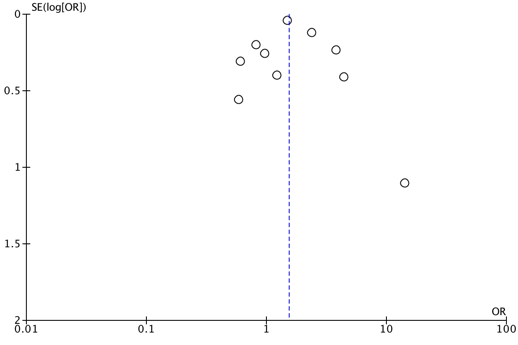 |
| Hematologic/Thromboembolic Complications | Immunologic Complications |
| 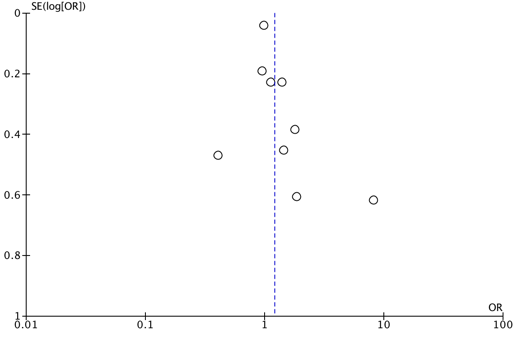 | 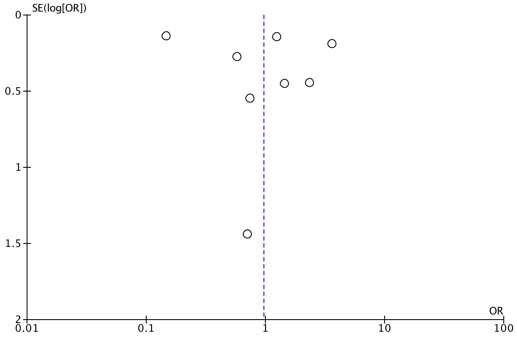 |
| Genitourinary Complications | Procedural Complications |
| 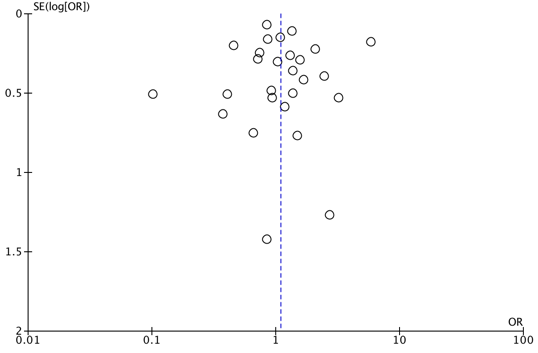 | 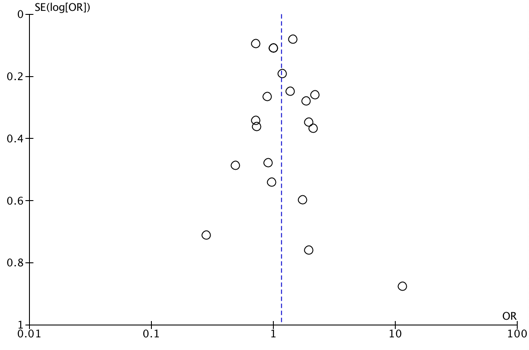 |
| <30 Day Mortality | >30 Day Mortality |
| 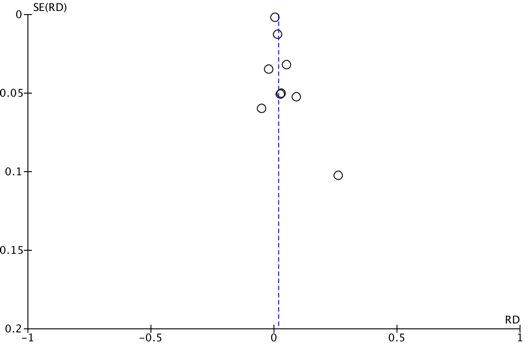 | 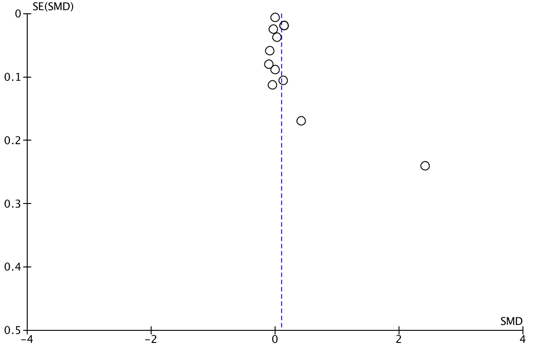 |
| Reoperation | Readmission |
| 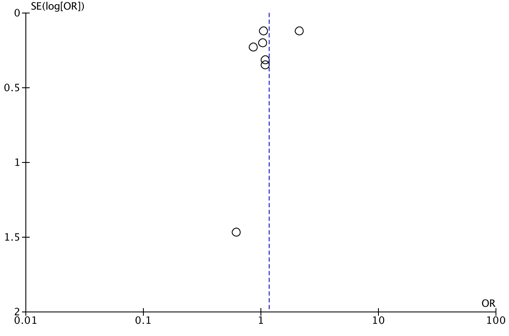 |  |
| Length of Hospital Stay |  |
