## Supplementary material for "Post-operative morbidity and mortality in Indigenous Peoples: A scoping review and meta-analysis": S3 Table

**S3 Table: Meta-Analyses of mortality, morbidity and hospital stay outcomes with associated subgroup analyses**

|  |  |  |  | Number of Studies (n) | Indigenous Patients  # Events/Total Population | Non-Indigenous Patients  # Events/Total Comparator | Odds Ratio (95%CI), p-value OR  SMD (95%CI), p-value* |
| --- | --- | --- | --- | --- | --- | --- | --- |
| Mortality |  |  |  |  |  |  |  |
|  | >30-Day Mortality | | | 20 | 1117/11825 | 33757/362441 | 1.16 (0.95-1,41), p=0.15 |
|  |  | Good Quality Studies | | 8 | 696/10396 | 3065/253977 | 1.03 (0.84-1.28), p=0.75 |
|  |  | Geographic Location | | | | |  |
|  |  |  | **Oceania** | **13** | **907/9626** | **2934/66461** | **1.29 (1.06-1.57), p=0.01** |
|  |  |  | Australia | 6 | 402/1403 | 856/4757 | 1.19 (0.93-1.51), p=0.17 |
|  |  |  | **New Zealand** | **7** | **505/8223** | **2078/61704** | **1.39 (1.01-1.92), p=0.04** |
|  |  |  | North America (USA) | 7 | 1117/11825 | 33757/362441 | 0.85 (0.58-1.24), p=0.39 |
|  |  | Surgical Specialty | |  |  |  |  |
|  |  |  | Cardiovascular Surgery | 9 | 214/898 | 1164/5474 | 1.33 (0.93-1.91), p=0.11 |
|  |  |  | General Surgery | 3 | 93/1447 | 1786/192080 | 0.94 (0.58-1.53), p=0.81 |
|  |  |  | Orthopedic Surgery | 3 | 47/7906 | 160/60188 | 1.31 (0.96-1.79), p=0.09 |
|  |  |  | Urology | 4 | 974/11204 | 4832/264678 | 1.03 (0.54-1.98), p=0.92 |
|  |  | Publication Period | |  |  |  |  |
|  |  |  | 2017 and later | 12 | 848/10525 | 3742/254185 | 1.17 (0.94-1.45), p=0.16 |
|  |  |  | Before 2017 | 8 | 269/1300 | 30015/108256 | 1.20 (0.79-1.81), p=0.39 |
|  | <30 Day Mortality | | | 28 | 773/49455 | 63579/7224799 | 1.20 (0.81-1.78), p=0.37 |
|  |  | Good Quality Studies | | 8 | 609/38910 | 50417/5808790 | 0.94 (0.66-1.35), p=0.75 |
|  |  | Geographic Location | |  |  |  |  |
|  |  |  | Oceania | 14 | 307/10900 | 5437/133247 | 1.03 (0.76-1.39), p=0.85 |
|  |  |  | Australia | 10 | 113/2829 | 4937/72272 | 0.94 (0.57-1.56), p=0.81 |
|  |  |  | New Zealand | 4 | 194/8071 | 500/60975 | 1.18 (0.87-1.59), p=0.28 |
|  |  |  | North America | 14 | 466/38555 | 58142/7091552 | 1.44 (0.71-2.92), p=0.31 |
|  |  |  | Canada | 3 | 21/694 | 506/12378 | 0.77 (0.49-1.20), p=0.24 |
|  |  |  | USA | 11 | 445/37861 | 57636/7079174 | 1.57 (0.70-3.54), p=0.27 |
|  |  | Surgical Specialty | |  |  |  |  |
|  |  |  | Cardiovascular Surgery | 16 | 387/23933 | 32086/2512201 | 1.08 (0.88-1.34), p=0.45 |
|  |  |  | General Surgery/Urology | 8 | 173/11976 | 30205/3915721 | 0.87 (0.35-2.17), p=0.76 |
|  |  |  | **Orthopedic Surgery** | **3** | **159/10754** | **619/319343** | **1.32 (1.08-1.61), p=0.006** |
|  |  | Publication Period | |  |  |  |  |
|  |  |  | 2017 and later | 15 | 529/35272 | 43819/5743979 | 1.17 (0.81-1.67), p=0.40 |
|  |  |  | Before 2017 | 13 | 244/14183 | 19760/1480820 | 1.25 (0.95-1.41), p=0.62 |
| Morbidity |  |  |  |  |  |  |  |
|  | **Surgical Infections** | | | **22** | **985/30427** | **66202/2840558** | **1.34 (1.12-2.59), p=0.001** |
|  |  | **Good Quality Studies** | | **11** | **818/23282** | **63494/2600149** | **1.36 (1.10-1.67), p=0.004** |
|  |  | Geographic Location | |  |  |  |  |
|  |  |  | Oceania | 12 | 79/2465 | 3850/102247 | 1.40 90.89-2.22), p=0.15 |
|  |  |  | Australia | 9 | 75/2209 | 3840/101560 | 1.55 (0.91-2.64), p=0.11 |
|  |  |  | New Zealand | 3 | 4/256 | 10/687 | 0.75 (0.24-2.41), p=0.64 |
|  |  |  | **North America** | **10** | **906/27962** | **62352/2738311** | **1.33 (1.10-1.60), p=0.003** |
|  |  |  | Canada | 3 | 58/703 | 828/12339 | 1.31 (0.99-1.74), p=0.06 |
|  |  |  | **USA** | **7** | **848/27259** | **61524/2725972** | **1.36 (1.06-1.74), p=0.01** |
|  |  | Surgical Specialty | |  |  |  |  |
|  |  |  | **Cardiovascular Surgery** | **9** | **739/25227** | **61702/2630841** | **1.27 (1.18-1.37), p<0.001** |
|  |  |  | General Surgery/Urology | 7 | 216/3499 | 4101/116061 | 1.37 (0.94-1.98), p=0.10 |
|  |  |  | Obstetrics & Gynecology | 3 | 16/1467 | 327/89475 | 1.51 (0.22-10.53), p=0.68 |
|  |  |  | Orthopedic Surgery | 2 | 5/59 | 3/75 | 2.09 (0.46-9.62), p=0.34 |
|  |  | Publication Period | |  |  |  |  |
|  |  |  | **2017 and later** | **12** | **204/6370** | **3290/163716** | **1.50 (1.16-1.95), p=0.002** |
|  |  |  | Before 2017 | 10 | 985/30427 | 66202/2840558 | 1.16 (0.90-1.49), p=0.24 |
|  | Systemic Infections | |  | 7 | 585/23623 | 63900/254509 | 1.01 (0.93-1.10), p=0.87 |
|  |  | Good Quality Studies | | 6 | 560/21468 | 63878/2547245 | 1.00 (0.92-1.09), p=0.94 |
|  |  | Geographic Location | |  |  |  |  |
|  |  |  | Oceania | 2 | 7/112 | 13/104 | 0.55 (0.21-1.43), p=0.22 |
|  |  |  | North America | 5 | 578/23511 | 63887/2549405 | 1.01 (0.93-1.10), p=0.78 |
|  |  | Surgical Specialty | |  |  |  |  |
|  |  |  | Cardiovascular Surgery | 2 | 522/20274 | 62117/2433705 | 0.68 90.14-3.29), p=0.63 |
|  |  |  | General Surgery/Urology | 5 | 585/23623 | 63900/2549509 | 0.98 (0.72-1.33), p=0.89 |
|  |  | Publication Period | |  |  |  |  |
|  |  |  | 2017 and later | 5 | 553/21398 | 63867/2547171 | 1.01 (0.92-1.10), p=0.88 |
|  |  |  | Before 2017 | 2 | 32/2225 | 33/2338 | 1.01 (0.58-1.75), p=0.98 |
|  | Cardiovascular Complications | |  | 11 | 110/4817 | 694/149223 | 1.29 (0.66-2.52), p=0.46 |
|  |  | Good Quality Studies | | 6 | 59/3333 | 191/92365 | 1.24 (0.78-1.95), p=0.37 |
|  |  | Geographic Location | |  |  |  |  |
|  |  |  | Oceania | 4 | 41/775 | 453/1843 | 0.97 (0.25-3.73), p=0.97 |
|  |  |  | North America | 7 | 69/4042 | 241/147380 | 1.54 (0.97-2.43), p=0.07 |
|  |  | Surgical Specialty | |  |  |  |  |
|  |  |  | Cardiovascular Surgery | 6 | 66/895 | 554/2625 | 1.31 (0.45-3.79), p=0.62 |
|  |  |  | General Surgery/Urology | 3 | 44/2910 | 110/87394 | 1.36 (0.53-3.46), p=0.52 |
|  |  | Publication Period | |  |  |  |  |
|  |  |  | 2017 and later | 7 | 35/2093 | 559/145751 | 0.98 (0.36-2.66), p=0.96 |
|  |  |  | Before 2017 | 4 | 75/2724 | 135/3472 | 1.65 (0.92-2.99), p=0.09 |
|  | Pulmonary Complications | |  | 11 | 602/24626 | 38627/2538049 | 1.42 (1.03-1.94), p=0.03 |
|  |  | Good Quality Studies | | 9 | 534/24121 | 38654/2537483 | 1.32 (0.92-1.90), p=0.13 |
|  |  | Geographic Location | |  |  |  |  |
|  |  |  | **Oceania** | **6** | **162/773** | **439/1354** | **1.64 (1.25-2.17), p<0.001** |
|  |  |  | North America | 5 | 440/23853 | 38188/2536695 | 1.13 (0.71-1.82), p=0.61 |
|  |  | Surgical Specialty | |  |  |  |  |
|  |  |  | **Cardiovascular Surgery** | **6** | **219/1277** | **1058/12876** | **1.85 (1.50-2.29), p<0.001** |
|  |  |  | General Surgery/Urology | 4 | 383/23174 | 37535/2521067 | 0.94 (0.84-1.04), p=0.24 |
|  |  | Publication Period | |  |  |  |  |
|  |  |  | 2017 and later | 6 | 394/21384 | 37592/2523210 | 1.22 (0.81-1.83), p=0.34 |
|  |  |  | **Before 2017** | **5** | **208/3242** | **1035/14839** | **1.64 (1.06-2.54), p=0.02** |
|  | Hematologic/Thromboembolic Complications | | | 14 | 438/8578 | 30591/742230 | 1.28 (0.97-1.70), p=0.08 |
|  |  | Good Quality Studies | | 9 | 318/6932 | 29651/705278 | 1.29 (0.91-1.84), p=0.15 |
|  |  | Geographic Location | |  |  |  |  |
|  |  |  | Oceania | 9 | 219/2127 | 1464/40021 | 1.21 (0.93-1.59), p=0.16 |
|  |  |  | North America | 5 | 219/6451 | 29127/702209 | 1.43 (0.70-2.90), p=0.32 |
|  |  | Surgical Specialty | |  |  |  |  |
|  |  |  | Cardiovascular Surgery | 7 | 189/2115 | 1493/40535 | 1.36 (0.99-1.87), p=0.06 |
|  |  |  | General Surgery/Urology | 4 | 243/6250 | 29046/697537 | 1.27 (0.64-2.52), p=0.50 |
|  |  | Publication Period | |  |  |  |  |
|  |  |  | 2017 and later | 8 | 293/5551 | 29931/735934 | 1.15 (0.83-1.60), p=0.41 |
|  |  |  | Before 2017 | 6 | 145/3027 | 660/6296 | 1.46 (0.86-2.49), p=0.16 |
|  | **Immunologic Complications** | |  | **10** | **1273/6037** | **27707/123872** | **1.53 (1.08-2.02), p=0.02** |
|  |  | Good Quality Studies | | 4 | 366/2780 | 3239/9141 | 1.47 (0.68-3.20), p=0.33 |
|  |  | Geographic Location | |  |  |  |  |
|  |  |  | **Oceania** | **3** | **299/508** | **3113/6360** | **2.35 (1.36-4.04), p=002** |
|  |  |  | North America | 7 | 974/5529 | 24594/117512 | 1.27 (0.80-2.02), p=0.32 |
|  |  | Surgical Specialty |  |  |  |  |  |
|  |  |  | Cardiovascular Surgery | 1 | 5/2155 | 9/2264 | 0.58 90.19-1.74), p=0.33 |
|  |  |  | **General Surgery/Urology** | **9** | **1268/3882** | **27698/121608** | **1.63 (1.14-2.33), p=0.007** |
|  |  | Publication Period | |  |  |  |  |
|  |  |  | **2017 and later** | **6** | **1193/3668** | **27201/120210** | **1.48 (1.00-2.20), p=0.05** |
|  |  |  | Before 2017 | 4 | 80/2369 | 506/3662 | 1.96 (0.64-6.00), p=0.24 |
|  | Genitourinary Complications | |  | 9 | 1097/6896 | 37423/206267 | 1.21 (0.92-1.60), p=0.18 |
|  |  | Good Quality Studies | | 5 | 148/3466 | 758/93967 | 1.15 (0.92-1.45), p=0.22 |
|  |  | Geographic Location | |  |  |  |  |
|  |  |  | Oceania | 5 | 130/1098 | 352/4319 | 1.43 (0.78-2.61), p=0.25 |
|  |  |  | North America | 4 | 967/5798 | 37071/201948 | 1.00 (0.78-2.61), p=0.90 |
|  |  | Surgical Specialty | |  |  |  |  |
|  |  |  | Cardiovascular Surgery | 5 | 130/1098 | 352/4319 | 1.43 (0.78-2.61), p=0.25 |
|  |  |  | General Surgery/Urology | 3 | 964/5623 | 37033/197842 | 0.99 (0.92-1.07), p=0.85 |
|  |  | Publication Period | |  |  |  | 1.14 (0.83-1.58), p=0.41 |
|  |  |  | 2017 and later | 6 | 995/4035 | 37182/201239 | 1.61 (0.74-3.51), p=0.23 |
|  |  |  | Before 2017 | 3 | 102/2861 | 241/5028 |  |
|  | Procedural Complications | |  | 8 | 183/11339 | 16429/1209334 | 0.96 (0.32-2.92), p=0.94 |
|  |  | Good Quality Studies | | 3 | 27/2914 | 449/87348 | 0.81 (0.41-1.62), p=0.56 |
|  |  | Geographic Location | |  |  |  |  |
|  |  |  | Oceania | 3 | 22/112 | 84/519 | 1.32 (0.56-3.14), p=0.53 |
|  |  |  | North America | 5 | 161/11227 | 16345/1208815 | 0.88(0.22-3.59), p=0.86 |
|  |  | Surgical Specialty | |  |  |  |  |
|  |  |  | General Surgery/Urology | 2 | 26/2872 | 448/87318 | 0.85 (0.35-2.09), p=0.73 |
|  |  |  | Orthopedic Surgery | 2 | 105/7518 | 15337/1066399 | 0.43 (0.05-3.97), p=0.45 |
| Hospital Stay Outcomes | | |  |  |  |  |  |
|  | Readmission |  |  | 7 | 299/3251 | 7797/163003 | 1.17 (0.83-1.64), p=0.37 |
|  |  | Good Quality Studies | | 3 | 134/1057 | 3298/89325 | 0.97 0.74-1.27), p=0.82 |
|  |  | Geographic Location | |  |  |  |  |
|  |  |  | Oceania | 2 | 90/860 | 3412/36568 | 1.05 (0.84-1.32), p=0.60 |
|  |  |  | North America | 5 | 209/2391 | 4385/126435 | 1.20 (0.73-1.98), p=0.46 |
|  |  | Surgical Specialty | |  |  |  |  |
|  |  |  | Cardiovascular Surgery | 2 | 90/860 | 3412/36568 | 1.05 (0.84-1.32), p=0.65 |
|  |  |  | General Surgery/Urology | 3 | 200/2209 | 4087/121275 | 1.26 (0.68-2.33), p=0.46 |
|  |  |  | Neurosurgery | 2 | 9/182 | 298/5160 | 1.05 (0.54-2.05), p=0.89 |
|  |  | Publication Period | |  |  |  |  |
|  |  |  | 2017 and later | 5 | 209/1842 | 6736/126503 | 1.01 (0.84-1.21), p=0.91 |
|  |  |  | Before 2017 | 2 | 90/1409 | 1061/36500 | 1.61 (0.84-3.12), p=0.15 |
|  | **Reoperation** |  |  | **9** | **142/3372** | **1795/149505** | **01.33 (1.02-1.74), p=0.04** |
|  |  | **Good Quality Studies** | | **5** | **94/617** | **159/2864** | **1.49 91.04-2.14), p=0.03** |
|  |  | Geographic Location | |  |  |  |  |
|  |  |  | **Oceania** | **7** | **105/732** | **249/4741** | **1.50 (1.05-2.13), p=0.03** |
|  |  |  | North America | 2 | 37/2640 | 1546/145764 | 1.13 (0.81-1.58), p=0.47 |
|  |  | Surgical Specialty | |  |  |  |  |
|  |  |  | Cardiovascular Surgery | 5 | 85/639 | 170/3270 | 1.47 (0.94-2.30), p=0.09 |
|  |  |  | General Surgery/Urology | 3 | 47/2661 | 1612/146117 | 1.23 (0.72-2.08), p=0.45 |
|  |  | Publication Period | |  |  |  |  |
|  |  |  | 2017 and later | 3 | 64/260 | 71/375 | 1.35 (0.61-2.97), p=0.46 |
|  |  |  | **Before 2017** | **6** | **78/3112** | **1724/149130** | **1.33 (1.02-1.74), p=0.03** |
|  | LOS |  |  | 12 | NA | NA | 0.09 (0.01-0.17), p=0.30 |
|  |  | **Good Quality Studies** | | **6** | **NA** | **NA** | **0.15 (0.02-0.29), p=0.02** |
|  |  | Geographic Location | |  |  |  |  |
|  |  |  | **Oceania** | **5** | **NA** | **NA** | **0.54 (-0.00-1.08), p=0.05** |
|  |  |  | North America | 7 | NA | NA | 0.03 (-0.04-0.10), p=0.43 |
|  |  | Surgical Specialty | |  |  |  |  |
|  |  |  | Cardiovascular Surgery | 5 | NA | NA | 0.52 (-0.03-1.07), p=0.06 |
|  |  |  | General Surgery/Urology | 4 | NA | NA | -0.00 (-0.01-0.01), p=0.97 |
|  |  |  | Orthopedic Surgery | 3 | NA | NA | 0.09 (-0.01-0.19), p=0.08 |
|  |  | Publication Period | |  |  |  |  |
|  |  |  | 2017 and later | 8 | NA | NA | 0.02 (-0.04-0.08), p=0.53 |
|  |  |  | **Before 2017** | **4** | **NA** | **NA** | **0.65 (0.14-1.16), p=0.01** |

Meta-analysis results of overall morbidity and mortality outcomes. Includes subgroup analysis by quality of study (good quality), geographic region (Oceania, North America), country, surgical specialty, and publication period (before 2017 or 2017 and later), if adequate data for analysis. Indigenous outcomes represent all reported Indigenous groups, including combining Pacific Islander and Native Hawaiian for relevant studies. General Surgery and Urology are combined when the only urologic surgeries include renal transplantation, which are done in some jurisdictions by general surgeons. Urology on its own combines renal transplantation with other urologic procedures. Significant results are presented in bold font. CI: Confidence Interval; ENT: Ears Nose Throat; LOS: Length of Hospital Stay; NA: Not Applicable; OR: Odds Ratio; SMD: Standard Mean Difference; USA: United States of America. *SMD only used for LOS continuous data
